# A quantitative multi-parameter mapping protocol standardized for clinical research in autoimmune neuroinflammatory diseases with white matter abnormalities

**DOI:** 10.1101/2024.10.07.24315008

**Authors:** Henri Trang, Tim J. Hartung, Qianlan Chen, Stefan Hetzer, Claudia Chien, Pia S. Sperber, Tanja Schmitz-Hübsch, Susanna Asseyer, Rebekka Rust, Darius Mewes, Lina Anderhalten, Michael Sy, Alexander U. Brandt, Carsten Finke, Friedemann Paul

## Abstract

Quantitative magnetic resonance imaging (qMRI) involves mapping microstructure in standardized units sensitive to histological properties and supplements conventional MRI, which relies on contrast weighted images where intensities have no biophysical meaning. While measuring tissue properties such as myelin, iron or water content is desired in a disease context, qMRI changes may typically reflect mixed influences from aging or pre-clinical degeneration. We used a fast multi-parameter mapping (MPM) protocol for clinical routine at 3T to reconstruct whole-brain quantitative maps of magnetization transfer saturation (MT), proton density (PD), longitudinal (R1), and transverse relaxation rate (R2*) with 1.6 mm isotropic resolution. We report reference MPM values from a healthy population with age and gender distributions typical of neuroimmunology studies in whole brain white matter (WM), T2-weighted WM hyperintensities, cortical grey matter and deep grey matter regions and present post-processing optimizations including integration of lesions and normalization of PD maps against cerebrospinal fluid (CSF) for standardized research in multiple sclerosis (MS) and related disorders. PD maps were affected by WM abnormalities in MS using WM calibration. The results acknowledge the impact of non-linear age effects on MPM and suggest using CSF calibration for future clinical application in autoimmune neuroinflammatory diseases with WM abnormalities.

## 1 Introduction

Quantitative magnetic resonance imaging involves mapping microstructure in standardized physical units containing information about the local tissue environment surrounding the protons, thereby enhancing comparability in time and space. Quantitative maps supplement conventional MRI, which relies on contrast weighted images where intensities have no biophysical meaning, in providing insight into biologically meaningful microstructural properties of the central nervous system at the mesoscopic scale ^1,2^. Research in quantitative relaxometry and magnetization transfer imaging has shown strong reproducibility and sensitivity, exhibiting a robust correlation with histological measurements and accepted metrics related to water ^3^, myelin ^4^, and iron content ^5^.

Standardized scanning protocols at 3T and tools to reconstruct parametric maps demonstrating multi-center reproducibility are readily available ^6,7^ such as the time-efficient multi-parameter mapping (MPM) protocol ^6^ consisting of 3D multi-echo fast low angle shot (FLASH) acquisitions. It allows for estimation of quantitative maps of proton density (PD), magnetization transfer saturation (MT), longitudinal relaxation rate (R1=1/T1), and effective transverse relaxation rate (R2*=1/T2*) facilitated by the open-source ‘hMRI toolbox’ ^8^, which includes spatial processing tailored for voxel-wise statistical analysis of quantitative cerebral MRI data.

Several studies have collected quantitative relaxometry, MT and PD maps of healthy brain tissue using the MPM protocol to obtain normative reference values ^7,9^. These studies highlighted the influence of normal aging on brain microstructure in previous quantitative MRI studies ^10,11^.

This study aimed to optimize the post-processing of a previously described MPM protocol based on standard manufacturer sequences with 1.6 mm isotropic voxel resolution ^12^ for disease-related research, i.e. autoimmune neuroinflammatory diseases with white matter lesions such as multiple sclerosis (MS) or neuromyelitis optica spectrum disorder (NMOSD). MS and NMOSD are immune-mediated inflammatory diseases of the central nervous system with overlapping clinical characteristics shown to predominantly affect women, given a markedly high female to male ratio in MS^13^ and NMOSD^14^. In a large healthy cohort with a gender distribution typical of neuroimmunology studies, we established reference values of MT, R1, PD and R2* in white matter (WM), T2-weighted (T2w) WM hyperintensities, cortical grey matter (CGM) and deep gray matter (DGM) regions. We compared WM and T2w WM hyperintensities values across MS patients and healthy controls. We standardized PD maps using reference values for water in cerebrospinal fluid (CSF) of the lateral ventricles ^15^ and compared them to maps scaled to 69% in WM to highlight the impact of focal and diffuse white matter damage between MS patients and healthy controls. We further evaluated MPM-derived parameters and their associations with age and sex to inform future studies that may require strategies to correct for confounding effects of these factors when using MPM in clinical research.

## 2 Materials and Methods

### 2.1 Subjects

#### 2.1.1 Informed Consent

The analysis was performed as part of the BERLimmun ^16^ (EA1/362/20, DRKS00026761), ViMS ^17^ (EA1/182/10) and CAMINO ^18^ (EA2/007/21) neuroimmunology studies, approved by the institutional ethics committee of our institution and conducted in accordance with the Declaration of Helsinki in its applicable version for the conduction of the study. All participants gave written informed consent.

#### 2.1.2 Study Population

Demographics are summarized in Table 1. From April 2015 to September 2022, we pooled healthy participants recruited from the 3 previous separate registries. For the control group, participants without history of neurological or psychiatric disorders (and without previous COVID-19 infection for the CAMINO cohort) were recruited in Germany. Additional inclusion criteria were the following: self-declared healthy, older than 18 years of age, an active health insurance, competent to give written informed consent. Exclusion criteria consisted of contraindication to MRI investigation at inclusion, pregnancy, disease hindering the conduct of the study or inability to cooperate. Initially, we collected scans from 78 healthy participants. We excluded one participant from analyses because scans were of poor image quality. In total 77 healthy controls, 60 (77.9%) women and 17 (22.1%) men, with age ranging from 20 to 75 years, with a mean [±standard deviation (SD)] of 42.1 ± 14.1 years, were included (14 from BERLimmun, 14 from ViMS, 49 from CAMINO). Additionally, 27 patients diagnosed with MS (18 women (67%), mean age 50 ± 9.9 years) were included from the ViMS study according to the revised McDonald diagnosis criteria ^19^.

**Table 1.**
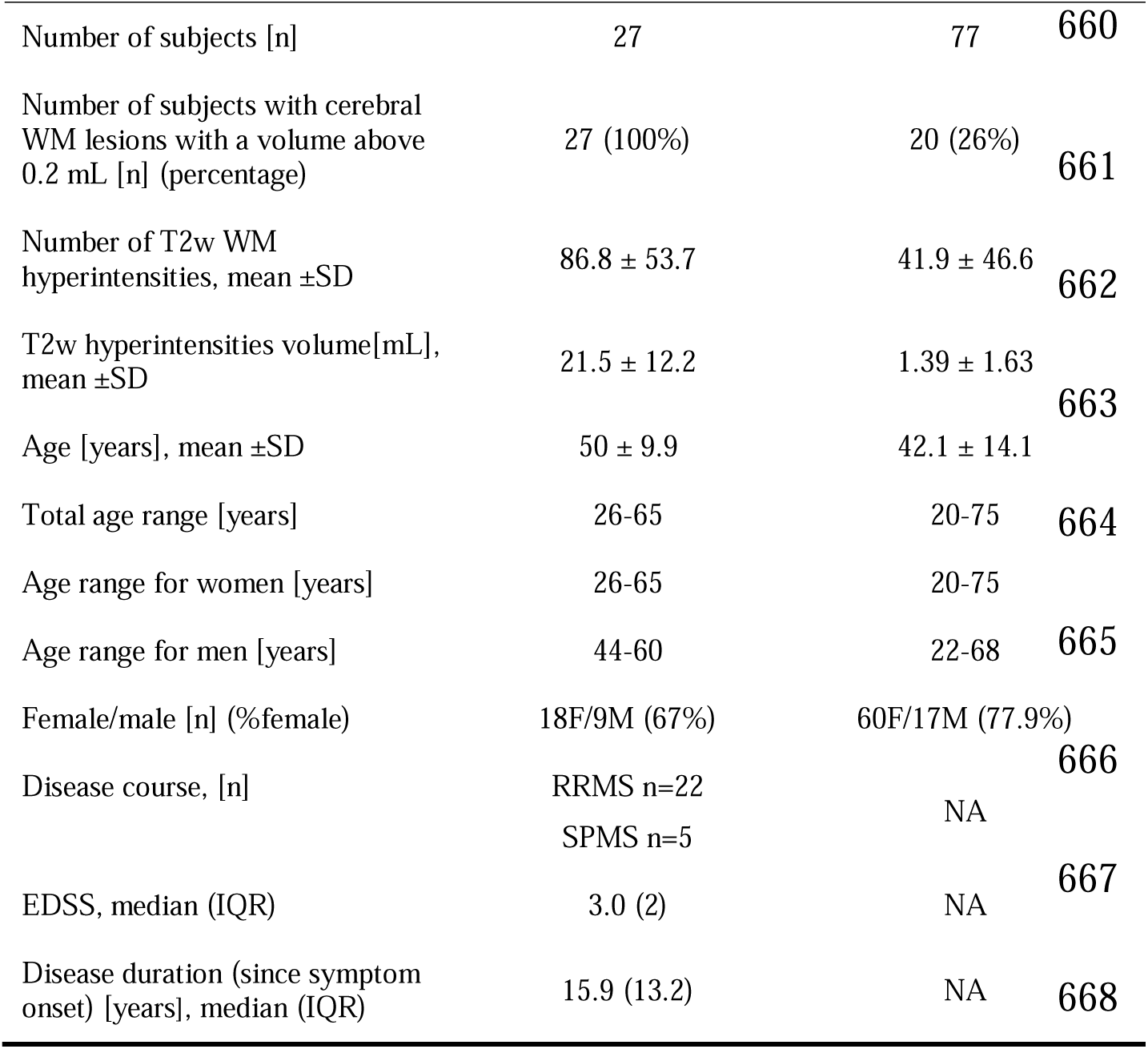
Demographics and clinical characteristics of the participants. MS patients were diagnosed as RRMS according to revised Mc Donald criteria^19^ and as SPMS according to Lublin et al. 2014^55^. Abbreviations: HC= healthy controls, F=female, M=male, WM=white matter, SD = standard deviation, IQR = interquartile range, RRMS = relapse-remitting multiple sclerosis, SPMS = secondary progressive multiple sclerosis, EDSS = Expanded Disability Status Scale.

### 2.2 MRI

#### 2.2.1 Acquisition

MRI scans were acquired on a single 3T MR scanner (Magnetom Prisma, Siemens Healthineers, Erlangen, Germany) using a 64-channel receive radiofrequency (RF) head-neck coil covering brain and cervical spinal cord. To maintain reproducibility across participants and time points, the acquisition protocol, and participant positioning remained identical to that detailed in a prior study, aside from updating the head coil ^12^. Briefly, the MPM sequence is 7 minutes in length, 1.6 mm isotropic resolution with three distinct 3D multi-echo fast low-angle shot (FLASH) gradient-echo acquisitions. For post-acquisition bias-field correction, a radiofrequency (RF) transmit (B1+) map was acquired during each session with an isotropic resolution of 4 mm from spin-echo/stimulated echo acquisitions utilizing a standard vendor sequence ^7^. MT-weighting was achieved by applying an off-resonance Gaussian pulse (500°, 10 ms, 1,200 Hz off-resonance, 192 Hz bandwidth) prior to non-selective excitation. In addition, the BERLimmun scan protocol included a structural T1-weighted (T1w) scan (3D MPRAGE, TR=1,900 ms, TE=2.55 ms, TI=900 ms, 0.8 mm isotropic resolution) and T2w fluid-attenuated inversion recovery (3D FLAIR, TR=6,000 ms, TE=388 ms, TI=2,100 ms, 0.8 mm isotropic resolution). The CAMINO scan protocol included a 3D-MPRAGE (1 mm isotropic resolution, TR=1900 ms, TE=2.22 ms, TI=2100 ms).

#### 2.2.2 Quantitative map reconstruction

We generated quantitative PD, MT, R1, and R2^∗^ maps utilizing MATLAB (MathWorks) with the hMRI toolbox ^8^ implemented within SPM12 (http://www.fil.ion.ucl.ac.uk/spm/software/spm12/). These maps were reconstructed given the PD-weighted (PDw), MT-weighted (MTw), and T1w echoes acquired through FLASH acquisitions and corrected for transmit and receive field inhomogeneities ^20^. Correction of Gibbs ringing artifacts ^21^ was performed prior to reconstruction of the quantitative maps, consisting in the removal of oscillatory patterns situated around tissue borders from all six echoes of the raw images (PDw, MTw, T1w). Motion degradation index for each of the raw averaged echo images (PDw, MTw, T1w) were obtained from the toolbox to identify scans with motion artifacts ^22^. Correlation of motion degradation index with age was assessed to evaluate the impact of motion on R2* variability with age (Sup. Fig. S1). Finally, given the use of an off-saturation MT pulse with a flip angle of 500°, we linearly rescaled MT maps to harmonize values for comparison to literature values obtained with a 220° flip angle, as recent evidence showed that a linear rescaling to harmonize MT maps across manufacturers effectively reduced the inter-site bias ^7^.

#### 2.2.3 Post-processing

T1-MPRAGE and T2-FLAIR images underwent bias-field correction using non-parametric non-uniform intensity normalization ^23^ and were subsequently reoriented to the Montreal Neurological Institute (MNI) standard reference space for further lesion delineation using FSL FLIRT (http://www.fmrib.ox.ac.uk/fsl).

#### 2.2.4 Segmentation

Two expert MRI technicians (15-17 years of experience) performed manual segmentation using ITK-SNAP (available at www.itksnap.org) of T2w hyperintense brain lesions on FLAIR images linearly co-registered to MPRAGE images ^24^. We subsequently refer to our segmentations as WM lesions (WML), rather than the general T2w-hyperintensities. We only included lesion masks with a WML mean volume above a pragmatic cutoff of 0.20 mL, corresponding to a Fazekas visual rating score of 1 ^25,26^. Generation of a brain mask and tissue segmentation of T1-MPRAGE images to obtain WM, CGM and DGM masks were achieved via FastSurfer ^27^. Lesion-filled WM masks were obtained by subtracting lesions from WM masks. Additionally, for each mask, voxels with T1 values higher than 4s within the tissue masks were removed for further correction of partial volume effect ^28^. All masks and structural images were then linearly co-registered via FSL FLIRT to native space using the T1w image as reference. Median parameter values were extracted from WM, CGM and several atlas-defined deep grey matter structures (thalamus, caudate nucleus, putamen, globus pallidus, hippocampus, amygdala, and nucleus accumbens) for each participant with both hemispheres summed.

#### 2.2.5 PD calibration

The PD map output from the hMRI toolbox was corrected for R2* by extrapolating the signal at TE=0ms and was originally calibrated as 69% water content in the WM ^8^. To demonstrate bias resulting from WM abnormalities in MS patients, PD maps calibrated using a whole WM mask and a lesion-filled WM mask were compared respectively in the whole WM region (including WML) and the normal appearing WM region free of WML (NAWM).

Subsequently, reconstructed PD maps from the hMRI toolbox were recalibrated as pure water (100 % reference) based on the median CSF signal in the lateral ventricles^28^, using a mask from the Harvard Oxford template distributed with FSL (Functional MRI of the Brain Software Library, http://www.fmrib.ox.ac.uk/fsl/) warped into subject native space. To reduce partial volume effects, lateral ventricles masks were eroded by 1 voxel then corrected by multiplying them with a CSF tissue mask obtained from the respective tissue probability map (threshold of 0.9). Finally, we excluded voxels with T1 values lower than 4s to obtain only voxels with pure water in CSF, based on the quantitative R1 map output from the reconstruction toolbox. To account for a potential bias introduced by CSF volume or T1 variability in CSF, we compared our method to a calibration selecting only the 100 voxels with the shortest T1 times above the 4s cutoff, therefore obtaining a mask with the same volume across participants over voxels with a small range of T1 above 4s. We calculated coefficients of variation (CoV) of the scaling factor, which is the multiplication factor obtained to scale each individual PD map by dividing 100 % by its respective median in CSF.

### 2.3 Statistical Analyses

#### 2.3.1.1 Descriptive statistics

Histogram analysis of brain tissues was first done to assess distribution of the entire dataset in WM, WML, CGM and DGM (Sup. Fig. S2). To get a better representation of a healthy population cross-sectional data for each brain region of interest (ROI), all subsequent analyses were performed on data free from respective outliers outside the ROI-specific 2^nd^-98^th^ percentile. Statistical analysis was conducted in R (R Core Team, https://www.r-project.org). Normality of data distribution was tested using a Shapiro-Wilk test. Median MPM metrics in each brain ROI were used for statistical analysis. Structural volume of the considered ROI was normalized by intracranial volume to obtain an adjusted volume and account for intracranial volume differences between men and women. Differences between WM and WML across MS patients and healthy controls were assessed using ANOVA and linear regression models adjusted for age and sex followed by post-hoc Tukey tests.

#### 2.3.1.2 Sex differences

Interaction between sex and age was tested before excluding the former variable as a possible covariate for MPM-derived parameters. Analysis of sex on median MPM parameter showed no difference between men and women, including normalized structure volume as an additional covariate in the linear regression model. Therefore, all regression models were fitted non-stratified, i.e. by combining data from both men and women.

#### 2.3.1.3 MPM age effects

For the assessment of the relationship between MPM-derived parameters and age, MS subjects were excluded. Linear and non-linear relationships between each MPM parameter and age were tested for every structure. A polynomial regression model was built for each tissue parameter with age, adding normalized volume and sex as covariables. Orthogonal polynomials were used to reduce multicollinearity effects of age predictors (e.g. covariance of age, age², age^3^). This was implemented in R using the “poly()” function from the “stats” package. Non-linear volume dependency with age was further assessed by exploring the significance of the quadratic term.

Additionally, visual inspection of MPM-by-age scatterplots with LOESS-fitted trend lines indicated that a 1-knot linear spline model could best fit the age-related distribution. We selected a cutoff of 55 years (≥55) for the spline, which represented a split at approximately the 80^th^ quantile. This is consistent with the upper age limit of most clinical drug trials in multiple sclerosis, as the confounding effect of vascular lesions and other comorbidities increases beyond this cut-off. Furthermore, diffusion tensor imaging studies revealed that age-related decline is more apparent in the fifth decade of life ^29^.

#### 2.3.1.4 Model selection

We compared the performance of linear, linear spline, quadratic, cubic and exponential models to choose the simplest best fitting model according to a likelihood ratio test in addition to Akaike information criterion (AIC) comparison. The simplest model was generally chosen when the likelihood ratio-test did not return a significant difference.

## 3 Results

### 3.1 PD calibration

We chose to standardize PD based on ventricular CSF (see Fig.1 for an illustration of the pipeline), since calibration methods using lesion-filled WM masks and whole WM masks in 27 MS patients resulted in different PD values in NAWM and WM (Fig.2a). In contrast, no difference was found between NAWM and WM in effective PD (without calibration) or when using CSF as reference (Fig.2b and 2c).

**Figure 1.**
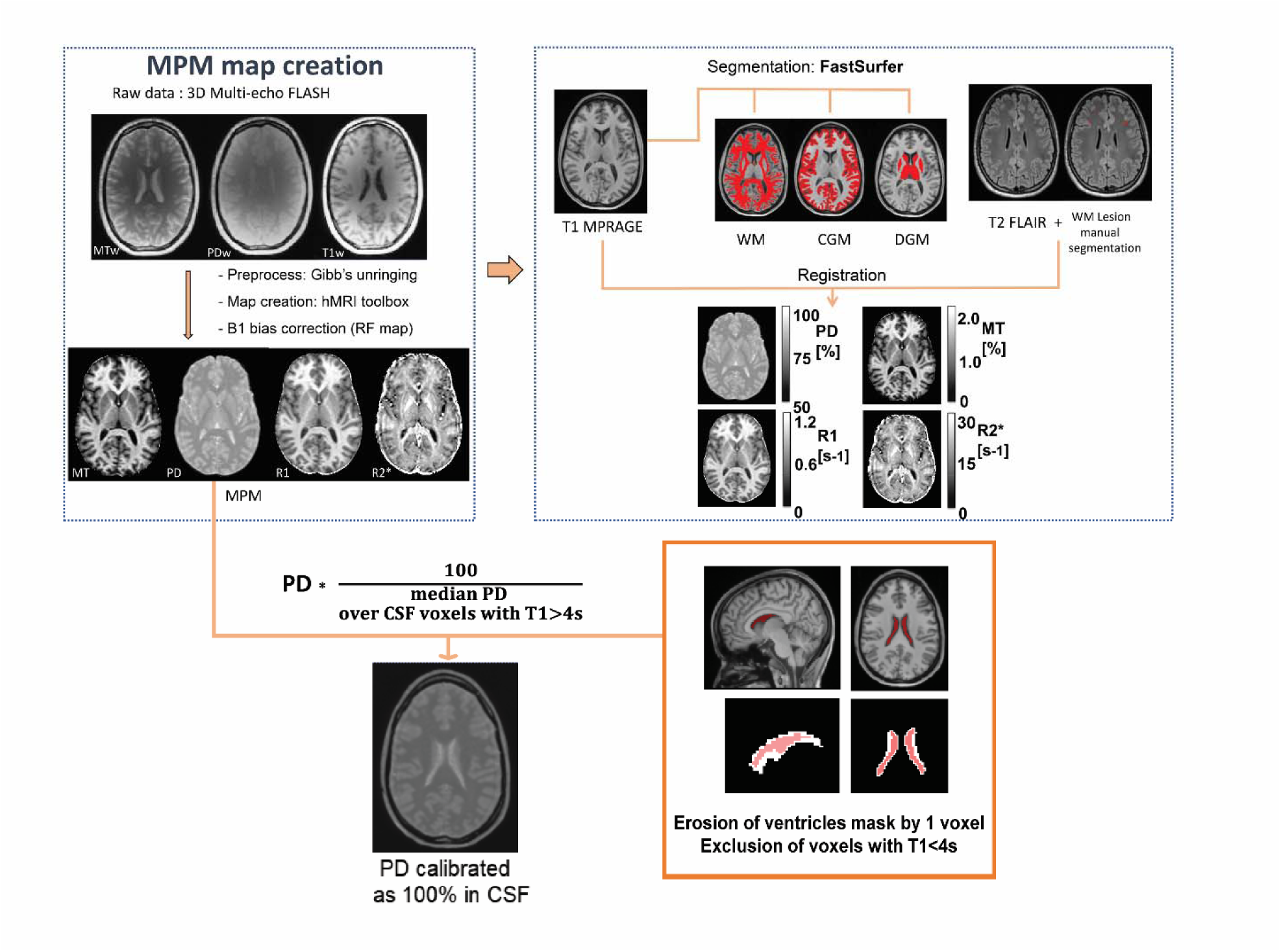
Graphical representation of the MPM pipeline. Raw PDw, MTw, T1w echoes were corrected for Gibb’s artifact before reconstruction with the hMRI toolbox. Receive field inhomogeneities were corrected using Unified Segmentation. T1-MPRAGE was segmented using Fastsurfer, a deep learning alternative to FreeSurfer, to obtain tissue masks for white matter (WM), cortical grey matter (CGM) and deep grey matter (DGM). White matter T2 hyperintensities were manually segmented from T2-FLAIR. All masks were then spatially registered to the quantitative maps. PD maps were calibrated as 100% in ventricular CSF. Voxels with T1<4s were excluded from the eroded lateral ventricles mask.

**Figure 2.**
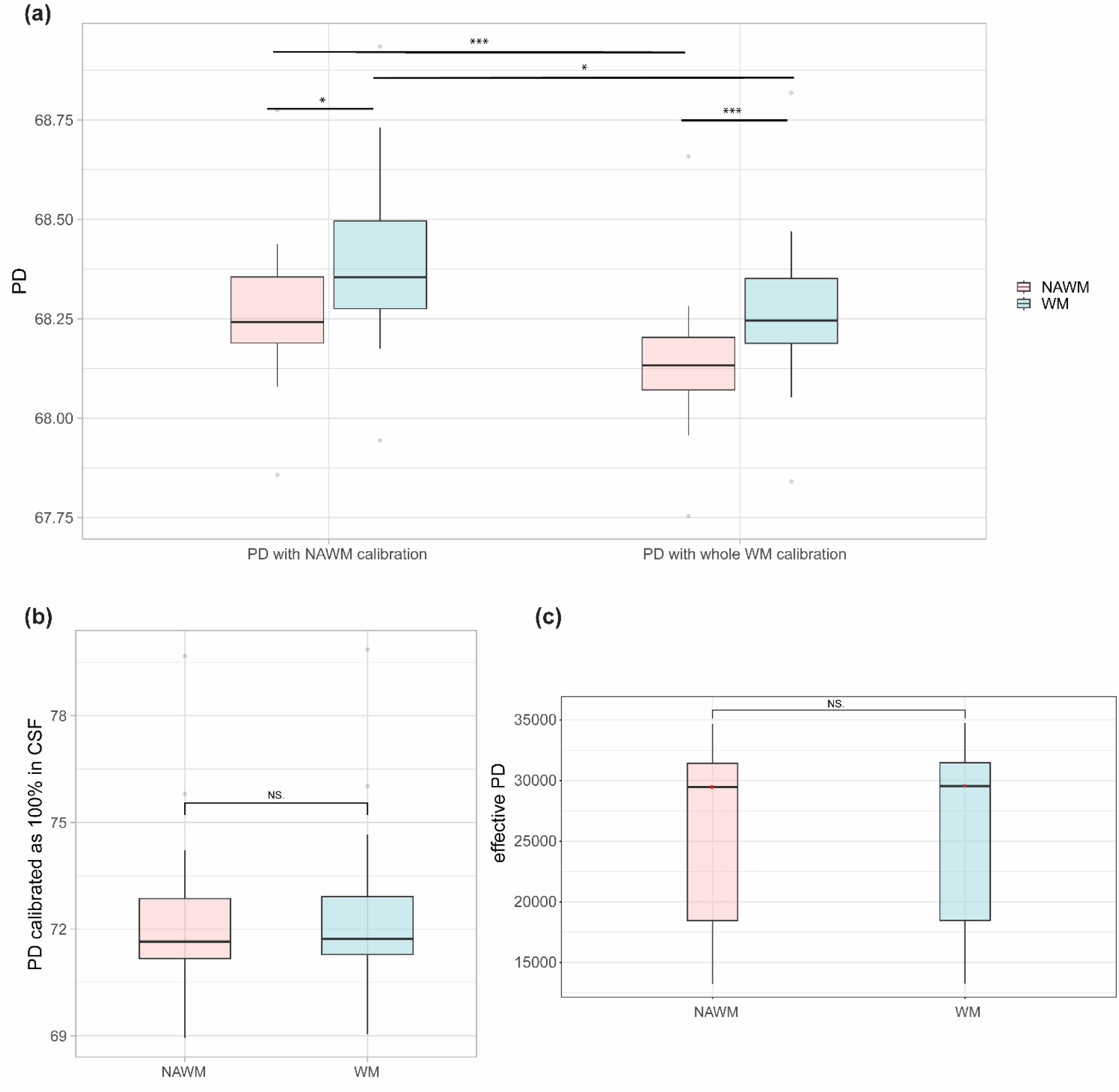
**(a)** PD comparison in whole white matter (WM) and normal-appearing white matter (NAWM) of 27 MS patients between calibration methods using lesion-filled white matter mask (PD with NAWM calibration) and whole white matter mask (PD with whole WM calibration). Significance levels associated to asterisks: p<0.05 (*), p<0.01 (**), p<0.001 (***). **(b)** Comparison of PD maps calibrated using CSF signal in normal appearing white matter (NAWM) and whole white matter (WM) regions of MS patients. Using CSF calibration, the mean difference between NAWM and WM is 0.1 p.u, which is of the same order of magnitude as in (a). Standard deviations are higher resulting in a higher inter-subject coefficient of variation. **(c)** Comparison of non-calibrated PD maps in normal appearing white matter (NAWM) and whole white matter (WM) regions of MS patients

Lateral ventricles masks had a median volume of 5685(7934) [median(IQR)] voxels or 23.3(32.5) mL. Exclusion of voxels with a T1 below 4s resulted in a median mask volume of 2683(4596) voxels or 10.99(18.82) mL. The average percentage of voxels remaining above this T1 threshold was 44.31% (mask voxel number ranging from 262 to 31396) with a range of T1 times from 4 to 13.4s. Median T1 across participants was 5.4s in this CSF mask. CoV of the scaling factor using CSF were 1.51%, 0.74% and 2.38% for respective T1 thresholds at 3s, 4s and 5s, compared to 37.79% for the calibration using WM. Using only the first 100 voxels with T1>4s in the mask resulted in a CoV of the scaling factor of 2.20%. Normalized lateral ventricles volume was correlated with age and scaling factor for the calibration using CSF, as well as scaling factor with age. However, there was no association between PD in the lateral ventricles and age (Sup. Fig. S3). After normalization with the CSF signal, mean intra-subject CoV of PD was 5.2 ± 0.33% in WM, 8.26 ± 0.58% in CGM and 4.84 ± 0.48% in DGM. Intra-subject variability remained identical to the WM-based calibration due to the linear rescaling of effective PD. SD of PD across subjects were higher in WM (1.27 vs 0.17), CGM (1.10 vs 0.68) and DGM (1.19 vs 0.69) with the normalization to CSF compared to calibration of PD against WM resulting in higher inter-subject CoV in WM (1.82% vs 0.24%), in CGM (1.40% vs 0.86%) and in DGM (1.54% vs 0.88%).

### 3.2 Healthy cohort reference values

Table 2 presents descriptive statistics of MPM measurements in WM, WML, CGM, DGM, thalamus, hippocampus and CSF. Data for caudate nucleus, putamen, globus pallidus, amygdala, nucleus accumbens are presented in Sup. Tables S1-2. Reported mean or median values across ROIs are in line with those reported in previous studies (Sup. Table S2).

**Table 2.** Descriptive statistics for all MPM and volume measurements in white matter (WM), white matter lesions (WML), cortical grey matter (CGM), deep grey matter (DGM), thalamus, hippocampus and CSF. Values are rounded at 2 decimals. Standard deviation over ROI voxels and intra-subject coefficient of variation are averaged across participants. Min - Max represent minimum and maximum of mean ROI values across participants. CSF values were extracted from the lateral ventricles excluding voxels with T1 values lower than 4s. Please note that PD values are scaled to 100 p.u. normalized by the median CSF value. MT values have been linearly rescaled to reference values of a MT pulse of 220° for comparison purpose to literature. Abbreviations: SD standard deviation, IQR interquartile range, CoV coefficient of variation, ROI region of interest, CSF cerebrospinal fluid

| Parameter |  | MT (%) |  |  |  |  |  |
| --- | --- | --- | --- | --- | --- | --- | --- |
| ROI | WM | WML | CGM | DGM | Thalamus | Hippocampus | CSF |
| Mean (SD) | 1.55 (0.05) | 1.24 (0.19) | 0.85 (0.02) | 1.05 (0.04) | 1.17 (0.05) | 0.83 (0.03) | 0.023 (0.007) |
| SD over voxels | 0.28 | 0.3 | 0.27 | 0.24 | 0.24 | 0.18 | 0.020 |
| Median (IQR) | 1.6 (0.06) | 1.26 (0.27) | 0.85 (0.03) | 1.03 (0.04) | 1.2 (0.07) | 0.82 (0.03) | 0.017 (0.008) |
| Intra-subject CoV (%) | 18.16 | 25.23 | 31.40 | 22.77 | 20.23 | 21.41 | 89.25 |
| Inter-subject CoV (%) | 3.21 | 15.28 | 2.78 | 3.43 | 4.13 | 4.12 | 30.01 |
| 2nd - 98th Percentile | 1.42 - 1.64 | 0.91 - 1.57 | 0.8 - 0.89 | 0.97 - 1.12 | 1.07 - 1.26 | 0.75 - 0.89 | 0.01 - 0.05 |
| Min - Max (of means) | 1.4 - 1.64 | 0.87 - 1.62 | 0.79 - 0.89 | 0.95 - 1.12 | 1.04 - 1.26 | 0.73 - 0.91 | 0.01 - 0.05 |
|  |  | PD (%) |  |  |  |  |  |
| ROI | WM | WML | CGM | DGM | Thalamus | Hippocampus | CSF |
| Mean (SD) | 69.97 (1.27) | 72.91 (2.94) | 78.73 (1.10) | 77.37 (1.19) | 76.53 (1.20) | 78.9 (1.27) | 100.72 (0.33) |
| SD over voxels | 3.64 | 4.63 | 6.5 | 3.75 | 4.11 | 3.24 | 4.55 |
| Median (IQR) | 69.08 (1.84) | 72.8 (4.11) | 78.81 (1.36) | 77.76 (2.05) | 76.61 (1.78) | 79.14 (1.72) | 100.14 (0.23) |
| Intra-subject CoV | 5.20 | 6.41 | 8.26 | 4.84 | 5.38 | 4.11 | 4.52 |
| Inter-subject CoV | 1.82 | 4.03 | 1.40 | 1.54 | 1.57 | 1.60 | 0.33 |
| 2nd - 98th Percentile | 67.67 - 72.22 | 67.13 - 77.48 | 76.32 - 80.76 | 75 - 79.26 | 74.15 - 78.77 | 76.47 - 80.82 | 100.27 - 101.85 |
| Min - Max | 66.87 - 72.53 | 66.24 - 77.56 | 76.29 - 81.17 | 74.03 - 79.32 | 73.78 - 79.09 | 75.27 - 80.93 | 100.04 - 101.98 |
|  |  | R1 (s<sup>-1</sup>) |  |  |  |  |  |
| ROI | WM | WML | CGM | DGM | Thalamus | Hippocampus | CSF |
| Mean (SD) | 0.91 (0.03) | 0.81 (0.08) | 0.62 (0.01) | 0.7 (0.02) | 0.73 (0.02) | 0.58 (0.02) | 0.185 (0.005) |
| SD over voxels | 0.12 | 0.13 | 0.1 | 0.11 | 0.09 | 0.07 | 0.016 |
| Median (IQR) | 0.92 (0.04) | 0.82 (0.16) | 0.61 (0.02) | 0.7 (0.03) | 0.73 (0.03) | 0.57 (0.02) | 0.186 (0.009) |
| Intra-subject CoV | 13.62 | 16.64 | 16.34 | 15.59 | 12.84 | 11.25 | 8.71 |
| Inter-subject CoV | 2.91 | 9.29 | 2.07 | 2.90 | 3.41 | 2.96 | 2.61 |
| 2nd - 98th Percentile | 0.85 - 0.96 | 0.7 - 0.94 | 0.59 - 0.64 | 0.67 - 0.74 | 0.67 - 0.77 | 0.55 - 0.62 | 0.17 - 0.19 |
| Min - Max | 0.83 - 0.96 | 0.7 - 0.95 | 0.58 - 0.64 | 0.66 - 0.74 | 0.65 - 0.78 | 0.54 - 0.62 | 0.14 - 0.20 |
|  |  | R2* (s<sup>-1</sup>) |  |  |  |  |  |
| ROI | WM | WML | CGM | DGM | Thalamus | Hippocampus | CSF |
| Mean (SD) | 21.29 (0.56) | 18.99 (4.11) | 18.01 (0.59) | 20.89 (1.07) | 20.23 (0.97) | 15.92 (0.78) | 2.23 (0.67) |
| SD over voxels | 4.23 | 5.48 | 9.19 | 6.44 | 3.57 | 3.9 | 2.06 |
| Median (IQR) | 21.06 (0.87) | 18.24 (2.27) | 16.8 (0.78) | 20.24 (1.13) | 20.37 (1.09) | 15.61(0.84) | 1.74 (0.52) |
| Intra-subject CoV | 19.87 | 25.69 | 51.03 | 30.66 | 17.67 | 24.44 | 90.37 |
| Inter-subject CoV | 2.64 | 21.61 | 3.30 | 5.11 | 4.80 | 4.92 | 30.03 |
| 2nd - 98th Percentile | 20.4 - 22.23 | 15.58 - 29.6 | 16.96 - 18.96 | 19.06 - 23.32 | 18.4 - 22.28 | 14.63 - 17.71 | 1.39 - 5.16 |
| Min - Max | 20.11 - 22.32 | 15.44 - 34.94 | 16.78 - 19.02 | 18.68 - 24.12 | 18.27 - 22.61 | 14.44 - 18.97 | 1.32- 5.98 |
|  |  | Volume (mL) |  |  |  |  |  |
| ROI | WM | WML | CGM | DGM | Thalamus | Hippocampus | CSF |
| Mean (SD) | 182.15 (9.52) | 1.39 (1.63) | 199.85 (11.37) | 15.61 (1.45) | 5 (0.45) | 1.05 (0.9) | 18.17 (20.35) |
| Mean ICV (mL) |  | 2890 (256.05) |  |  |  |  |  |

Histograms (Fig.3) show clearly distinct peaks and normal distributions of MPM median values for WM, WML, CGM and DGM. Detailed distribution density of median parameter values in DGM structures is shown in Figure 4a. The pallidum stands out among the deep grey matter structures with higher MT, R1 and R2* and lower PD (Fig.4a). This is further illustrated in Figure 4b displaying a brain slice sampled from each population averaged parameter map.

**Figure 3.**
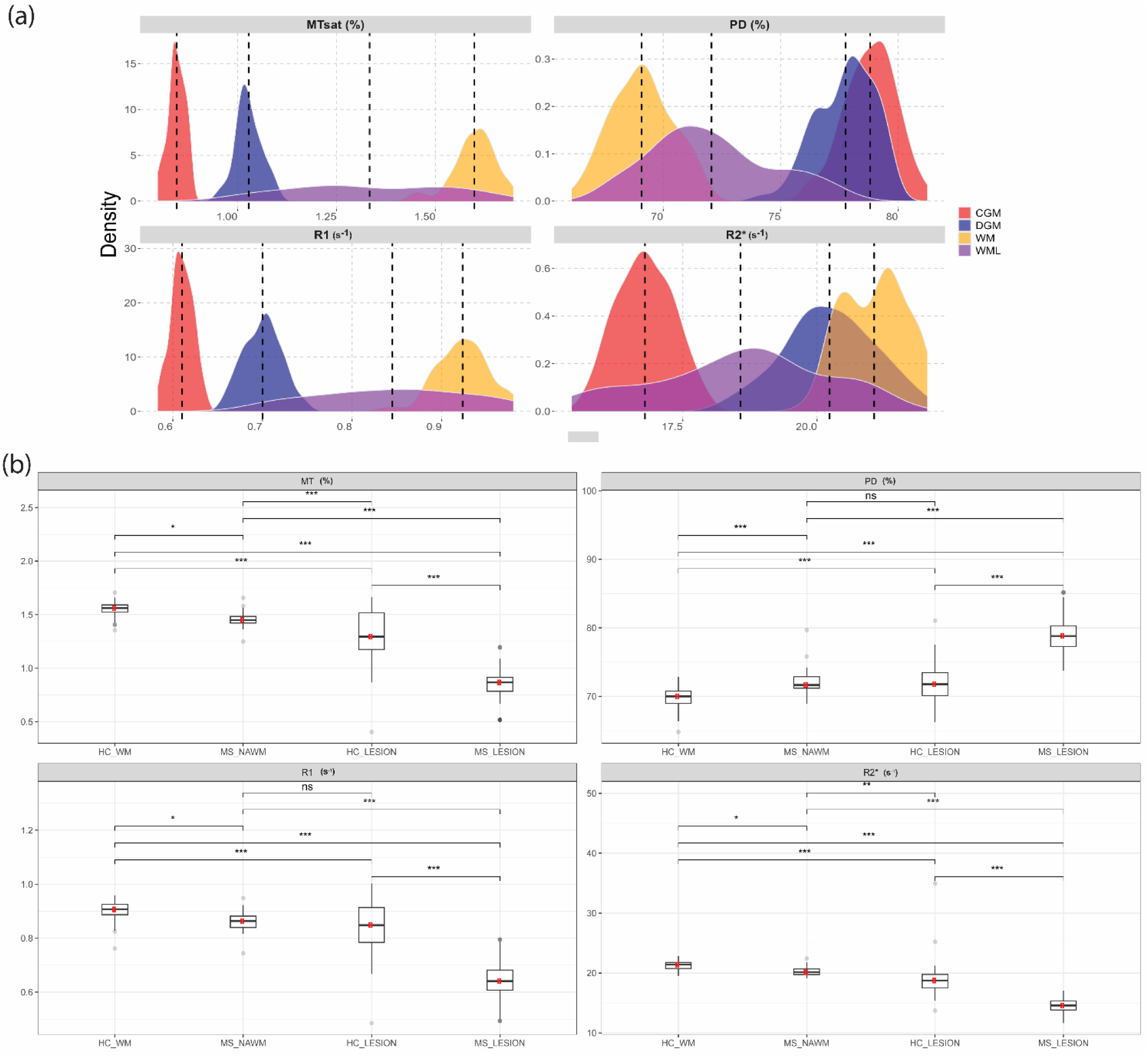
**(a)** Histograms of median MPM values distribution across healthy participants in white matter (WM, yellow), white matter lesions (WML, purple), cortical grey matter (CGM, red), deep grey matter (DGM, blue). Dashed lines indicate respective median. For each tissue class except WML, outliers outside the 2-98^th^ percentile were removed. WML only included median values of healthy participants with mean volume higher than 0.2mL. **(b)** MPM comparison of white matter lesions in MS patients (MS_LESION) and HC T2w white matter hyperintensities (HC_LESION) against healthy white matter (HC_WM) of healthy controls and normal appearing white matter of MS patients (MS_NAWM, free of lesions). Significance levels associated to asterisks: p<0.05 (*), p<0.01 (**), p<0.001 (***).

**Figure 4.**
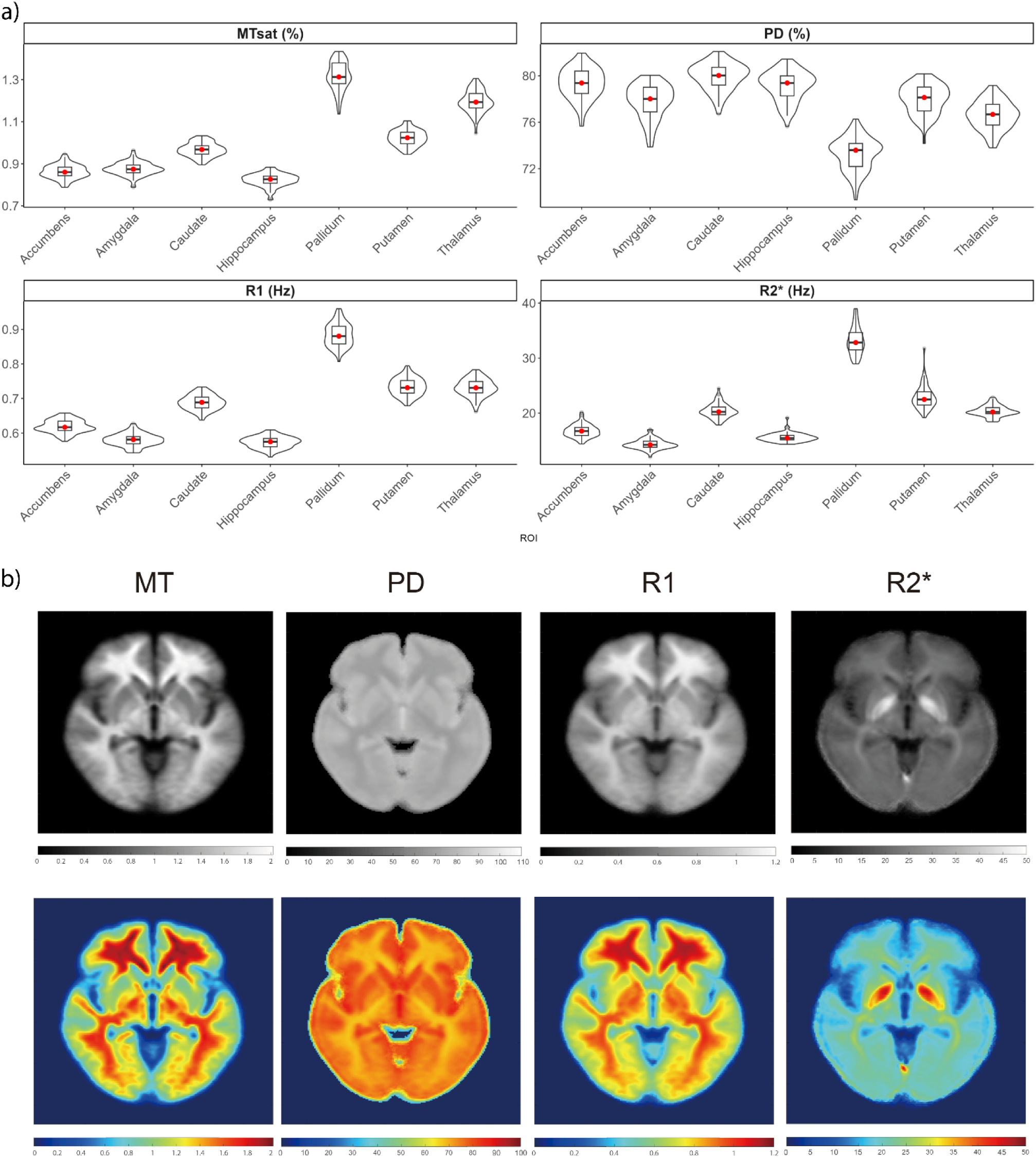
**(a)** Boxplots and density distribution comparison of median MT, PD, R1 and R2* values in thalamus, caudate, globus pallidus (Pallidum), putamen, amygdala, hippocampus, nucleus accumbens (Accumbens). **(b)** Greyscale and RGB-colored slice examples of population averaged quantitative maps showing globus pallidus caudate putamen and thalamus. In particular, the globus pallidus shows higher R2*, R1, MT and lower PD values.

### 3.3 White matter lesions

Fifty-two out of 77 healthy participants (67.5%) had at least one T2w WMH segmented as lesion (single lesion volume > 0.01 mL) and 20 (26%) participants (mean age 50.5±14y, 6 (30% male)) had a mean WML volume above the pre-defined cutoff of 0.2 mL. Mean (SD) number of lesions was 41.9 (46.6) and mean WML volume was 1.39 (1.63) mL (Table 1 and 2). MPM metrics and lesion volume scatterplots against age are shown in Sup. Fig. S4.

In healthy participants, compared to normal appearing WM, MPM values in WML were significantly reduced for MT (t=-7.53, 95% CI =[-0.43, -0.24], p < 0.001), R1 (t=-5.83, 95% CI =[-0.14, -0.07], p < 0.001) and R2*(t=-8.05, 95% CI =[-3.56, -2.09], p < 0.001) and significantly increased for PD (t=5.51, 95% CI =[2.31, 5.12], p < 0.001). In addition, WML showed a substantially wider range of MPM values across all parameters, compared to healthy WM (Fig.3).

MS patients had a mean T2w WMH volume of 21.5 (12.2) mL ranging from 10.3 mL to 58.2 mL across patients and a mean number of lesions of 86.8 (53.7). MS-WML had increased PD and reduced MT, R1 and R2* compared to MS-NAWM, HC-WM and HC-WML (Fig.3b).

### 3.4 MPM age related changes

As part of our objective to explore the impact of brain aging on the quantitative maps, Table 3 compiles effect sizes from the regression models, including 95% confidence intervals. Only models revealing a significant association between parameter and age in WM, CGM, DGM, thalamus and hippocampus are presented in Table 3 and outcomes of model selection are summarized in Sup. Fig. S5. For data visualization, scatterplots with the trajectory of the curve fitting in the various ROIs are attached in Fig.5 and Sup. Fig. S6.

**Figure 5.**
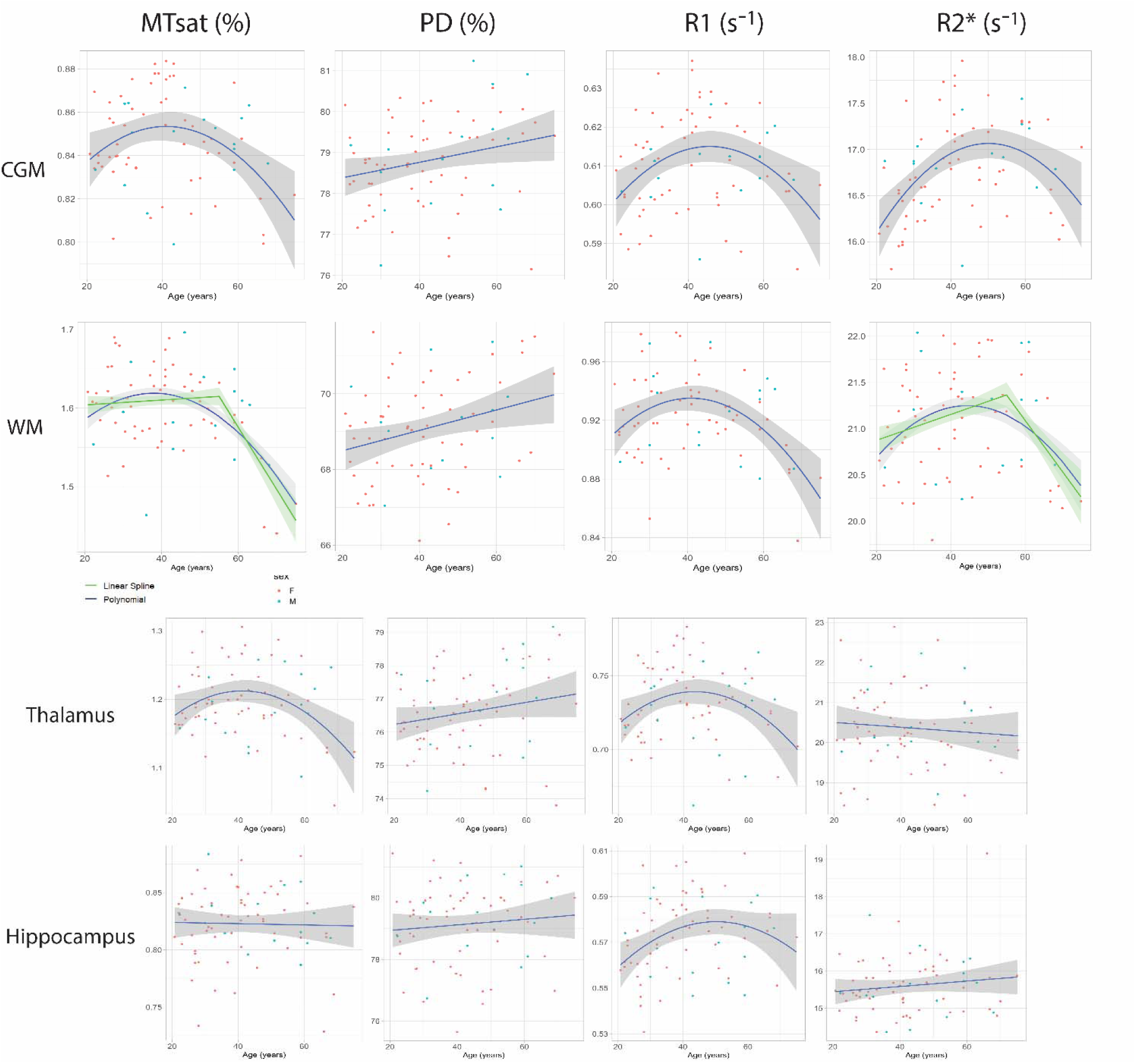
Scatterplots and fitted trajectories (blue) for the described ROIs. Green curve shows the linear spline with a cut-off of 55y when it performed better than the other models. Orange curve displays the cubic model which performed equally well but not significantly better than a linear regression in caudate nucleus and putamen. Red and green dots represent women and men respectively.

**Table 3.**
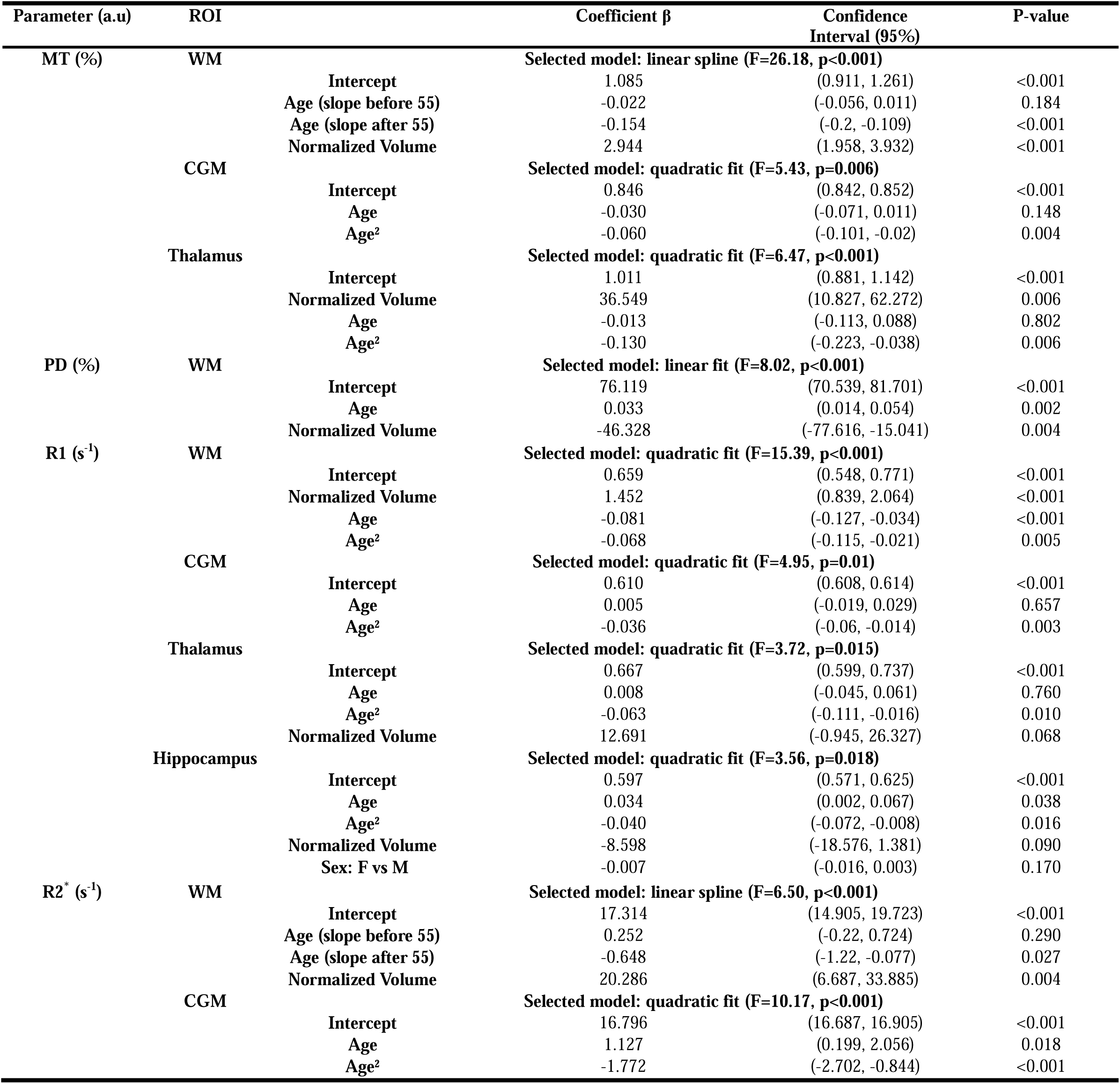
MPM selected models in white matter (WM), cortical grey matter (CGM), thalamus and hippocampus. Normalized volume was calculated as structural volume divided by intra-cranial volume. Age coefficients estimates are given in respective parameter unit per year (a.u/y).

Regression of MT, R1 and R2* showed a non-linear association with age in the quadratic term in both WM and CGM. In the thalamus, fitting MT and R1 with a quadratic function against age showed significant association, both described by a slow increase from 20 to 40y followed by a decline after 50y. In the hippocampus only R1 presented a quadratic evolution with age indicating a slow increase from 20 to 50y followed by a decline after 60y.

In WM, a linear spline performed better than a quadratic fit in explaining changes in MT and R2* with age. Summarizing the results of the model selection in WM (Table 3), MT showed an average decrease of -0.154%/year (p <0.001) after 55y, R2* a decrease of -0.648s^-1^/year (p <0.001) after 55y and PD increased linearly by 0.033%/year (p=0.002). R1 revealed a quadratic association with age (p=0.005) with a slow increase over 20-40y followed by a decline after 50y. Remarkably, there was no age association in both MT and R2* in WM between 20y and 55y.

In CGM, selected models were quadratic for MT, R1 and R2* and linear for PD. In the thalamus, a quadratic fit was selected for MT and R1 and in the hippocampus R1 also showed a quadratic trajectory with age.

Finally, there was no non-linear volume dependency with age across ROIs when testing the quadratic term, although correlation was present (Sup. Table S3).

## 4 Discussion

In this study we present an acquisition and post-processing protocol for a fast quantitative multi-parameter mapping of the brain with inclusion of T2w white matter hyperintensities and calibration of proton density using CSF. We standardize the reconstructed quantitative maps for future clinical application in demyelinating diseases and recommend calibrating PD maps as pure water in CSF to avoid bias introduced by pathology. We report reference MPM brain data for 77 healthy subjects of Caucasian ethnicity aged between 20 and 75 years and discuss age-dependence of the MPM parameters.

### 4.1 PD calibration

PD maps using the standard approach from the hMRI toolbox reconstruction are calibrated to 69% in WM which may lead to an underestimation of its reported variability ^7^. In addition, pathologic changes of PD in WM may not be reflected, especially when considering its possible use in diseases with WM abnormalities, e.g. multiple sclerosis. Using CSF-based calibration avoids these problems, as CSF does not contain relevant amounts of macromolecules or paramagnetic ions and the magnetic properties of CSF are largely unaffected by most neurological disorders ^30^.

We found that both diffuse and focal WM abnormalities affected the WM-based calibration. On one hand, focal lesions led to a slight underestimation of PD values. On the other hand, diffuse WM damage might also impact the calibration resulting in slight PD differences (∼0.1-0.2 p.u) between NAWM and WM while there was no difference in effective PD or when using CSF as reference. In such a ROI analysis, the impact from WM lesions may be occulted by the relative volume difference to whole white matter across patients. However, at the single subject level, their effect may be amplified. Therefore, we recalibrated PD as 100% in CSF, so that inter-subject variability in CSF would be negligible ^28^. To ensure that calibration is only based on voxels containing CSF and to avoid partial volume effects, we excluded voxels with T1 lower than 4s from the ventricle masks, which is an appropriate threshold given that the model estimated T1 of free water in brain tissue is 3.7s ^31^. Although T1 of CSF is known to be independent of field strength ^32^, reported values in literature show some variability ranging from 4s to 5s. The CoV of the scaling factor did not vary substantially between the usage of different T1 cut-offs from 3s to 5s. It was dramatically lower using CSF (0.74% vs 37.79%). We effectively scale all voxels based on the assumption that CSF has the highest water fraction. One could either assume that the highest water fractions might be capped by the maximum intensity measured by the scanner or that biological variability in CSF is of lower order of magnitude.

Inter-subject variability is higher than in a previous study which used the conventional WM-based calibration, while intra-subject CoV is in the same order of magnitude ^12^. However, our current sample has a wider age range (42.1±14.1 vs 35±7y) than the study by Cooper and colleagues. Given the variability of ventricles sizes, the resulting inter-subject CoV is higher compared to normalization to a larger homogeneous WM region. Nonetheless, taking into consideration the variation in ventricular volume in the linear models, both linear association of PD with age and CoV remained similar whether we used the ventricles mask or a fixed set of 100 voxels within it. Although correlation exists between ventricular volume and age, as well as between ventricular volume and scaling factor, the scaling factor was not correlated to age. This means that although age variability in ventricular volume introduces variability in the PD scaling factor, the resulting inter-subject variability remains low compared to inter-subject variability from WM. An alternative approach would be to obtain a scaling factor with the use of an external reference such as a phantom to benefit from a stable and homogeneous volume ^33^. However, due to a lack of practicality, it is hardly considered for use in a clinical setting.

Finally, PD quantification methods may differ on the determination of the receiver sensitivity profile and values reported here are standardized for the method of quantitative B1 mapping ^34^. Volz et al. pointed out that physiological bias might be smoothed out and recommended to proceed carefully in the presence of pathology which may cause segmentation algorithms to fail or wherein the relationship between T1 and effective PD may be locally distorted ^15^.

### 4.2 Age-related effects

We observed a non-linear age dependency of MT, R1 and R2* across various ROIs. Age effects could be best modelled linearly for PD and we found arguably a reduced impact of non-linear effects between 20y and 55y for both MT and R2* in WM. Our findings corroborate MPM studies on brain aging which reported negative correlation between age and MT across the cortex along positive correlations between age and R2* in the basal ganglia ^9–11,35^.

A quadratic model provided the most accurate representation of the non-linear relationship between MT and age, illustrating the U-shaped pattern in myelination over lifespan, consistent with myelin-driven changes in volume and MRI contrast across the cortex ^36^. Quantitative R1 has also shown a quadratic trajectory against age indicative of region-specific myelin maturation stabilizing into middle age followed by degeneration ^37^. Our results indicate that age-related MT and R1 changes were generally coincident, both sensitive to tissue myelin ^35^. However, R1 is less sensitive to myelin and reflects several physiological processes which can occur simultaneously, as modelled by its linear dependency on free water, myelin, macromolecules or iron assuming a mono-exponential decay ^31^.

Except for its linear increase in WM, we did not find any age-related association to PD in the investigated ROIs. Across the cortex, this is consistent with Seiler et al. who reported that global cortical PD did not show a significant correlation with age ^38^. Looking at interregional differences they detected in the temporal and occipital lobes a positive association with age. Filo et al. did not find differences in macromolecular tissue volume (MTV=1-PD, non-water tissue fraction) corrected for R2* in the frontal cortex, hippocampus, amygdala and frontal cortex between young adults and older adults ^39^. Overall, PD as a surrogate for water may be less sensitive to age-related changes of tissues, although values extracted from a single ROI do not allow generalization of the results to the whole cortex because of regional heterogeneity.

Linear and polynomial fits of R2* versus age performed significantly better on our data across ROIs than exponential saturation functions as the latter could not keep up with the increase in R2* and its variability in older participants. Yet early exponential growth in the putamen and caudate nucleus was noticed (Sup. Fig. S7) following the putative steep iron increase from early childhood to adulthood, in line with global cubic fits for the caudate, putamen, and globus pallidus reported in another study ^40^. Pallidal calcification was often seen in our older healthy participants, which we considered as a normal aging phenomenon contributing to the R2* inter-subject variability in the basal ganglia as hyperintensities may originate from the presence of minerals such as calcium and zinc especially around the globus pallidus ^41^. Although age-related increase in R2* has also been reported for the hippocampus ^42^ we did not observe a dependency of R2* with age in the hippocampus and thalamus, what may be a result of structural differences in rates of myelination or iron accumulation ^41^, or due to volume shrinkage impacting iron concentration.

Interestingly, we noticed a concurrence between R1, MT and R2* in WM and CGM owing to their sensitivity to macromolecular, myelin, iron and water content ^43^. Tissue areas rich in iron often co-localize with regions of elevated myelin content ^44^, owing to the role of iron in myelin synthesis and homeostasis, or the high iron concentration within glial cells ^45^, adding to the dependence of R2* on the orientation of WM fibers with respect to the magnetic field ^46^.

Finally, increased variability with age in R2* may partly be explained by noisier measurements given the sensitivity to motion inherent to the multi-echo FLASH acquisition. As shown by the correlation between age and motion degradation index, motion may also be a predictor as the ability to remain still in the scanner may worsen with age. Specifically, head motion extends to the noise level of relaxometry estimates derived from the raw echoes quantified by the variability of R2* in WM ^47^.

### 4.3 Implications for future research and clinical practice

Strengths of this study include using manufacturer sequences, which allow ready implementation on standard scanners. Absolute deviations in mean or median between our MPM values across ROIs and those reported in literature were in the same range as differences between previous studies highlighting the reproducibility of quantitative MPM ^6,7^. The prevalence and severity of WML tend to rise with age with a majority of non-demented people aged above 60 exhibiting cerebral lesions ^48^.

In consideration of future research implicating WM lesions and abnormalities in multiple sclerosis and related disorders, we aimed to include them in the general pipeline for lesion-filling and to improve PD quantification, as i) the presence of lesions or enlarged ventricles may cause segmentation to fail and ii) global effects observed in WM might transfer to lesion-specific localized effects. Comparing WM and WML across MS patients and healthy participants, T2w WM hyperintensities showed decreased MT, R1 and R2* and increased PD compared to healthy WM and NAWM of MS indicating more pronounced focal damage and structural loss. MPM is also sensitive to diffuse white matter pathology as NAWM which appears unaffected on conventional MRI can be differentiated from healthy WM ^49^. In particular, demyelination, axonal degeneration, inflammation, gliosis and edema are exacerbated in MS plaques resulting in higher discrepancy in MPM values from healthy WM and WML, corroborating findings from other studies using quantitative relaxometry, MT imaging, and diffusion MRI ^50^. However, MS lesions are heterogeneous and present varying degrees of degeneration, de/re-myelination and inflammation, thus discriminating specific lesion types remains to be explored ^51,52^.

Lastly, we hypothesized that MPM measurements might demonstrate more consistency in younger healthy individuals while displaying greater variability in older populations to acknowledge the impact of pre-clinical degeneration. Biological age may contribute most to the inter-subject variability and even be the strongest predictor for pathophysiological changes. Early and late nonlinear age-dependence have been observed in the lifespan trajectories of quantitative parameters, with distinct patterns in MT mimicking the inverted U-shaped trajectory of human brain myelination ^36^ and in R2* analogous to the exponential cerebral increase of non-heme iron ^53^. Yet, this relationship may be altered in disease. In multi-center or randomized clinical trials, due to discrepancy in age distribution of unmatched cohorts, including age as a linear predictor may be inadequate if one aims to fully capture the true age-related variability when manipulating biomarkers. It may be recommended to check and correct for non-linear age effects by fitting the response with age as an independent parameter. For example after fitting with an exponential saturation function, Ropele et al assessed inter-subject and inter-scanner variability of R2* and attributed large R2* variations to age suggestive of iron accumulation while scanner differences had a low impact ^54^.

### 4.4 Limitations

Although the sample size is small for robust non-linear models of age-related effects, we report healthy population MPM data which can serve as control data and such studies are scarce given the novelty of the protocol. Longitudinal studies are however superior to assess chronological pathological changes and reduce bias due to the large interindividual variability. Another liability is that the age and sex distributions of the recruited participants resemble those of typical cohorts of autoimmune neuroinflammatory diseases with a strong preponderance of women. This may however become a strength when studying such clinical populations. Indeed, our pipeline considered the inclusion of WML with MPM and is readily available for upcoming research in diseases like multiple sclerosis and related disorders. Consequently, the recruited population did not permit to have a good representation of early and late developmental changes occurring in the brain to effectively explore the impact of aging on quantitative maps and associate it to decline in cognition or motor function. Moreover, this study was done on a white population, so our results are not necessarily applicable to other ethnicities. In this study, no visual rating of the WML was attempted as they mostly served to establish the pipeline and will be further discussed and investigated in a following study to discriminate MS specific lesions from the WM lesions described in healthy participants which are likely microangiopathic. Future developments should target improvements in both sensitivity and specificity of MRI biomarkers, as well as clinical applicability with regards to disease models.

## 5 Conclusion

In conclusion, we present a fast quantitative MPM pipeline at 1.6 mm isotropic resolution, which can be readily used in a clinical protocol based on manufacturer sequences, along post-processing methods including standardization of PD maps and healthy brain data acquired with it. The protocol is anticipated to possess a higher sensitivity in identifying pathological alterations in future applications in disease ^12^. Importantly, previous studies ^6,7^, as well as the current study provide essential reference values and contribute datasets to assist clinical researchers in conducting thorough power analyses and report effect sizes that carry significance for future investigations in the context of clinical studies.

## Acknowledgments

This work was supported by the Berlin Center for Advanced Neuroimaging, by iNAMES - MDC - Weizmann - Helmholtz International Research School for Imaging and Data Science from NAno to MESo, the German Research Foundation (DFG) and the Open Access Publication Fund of Charité-Universitätsmedizin Berlin. For the studies ViMS and BERLimmun, we acknowledge institutional support by NCRC - Neuroscience Clinical Research Center funded by the Deutsche Forschungsgemeinschaft (DFG, German Research Foundation) under Germany‘s Excellence Strategy – EXC-2049–390688087 and Charité-BIH. We are thankful to our MR technicians, Cynthia Kraut and Susan Pikol, for their assistance in data acquisition.

## Author contributions

H.T: Formal analysis, Data curation, Methodology, Writing - original draft;

T.H: Formal analysis, Writing - review & editing;

Q.C.: Formal analysis, - review & editing; S.H: Methodology - review & editing;

C.C: Data curation, review & editing;

P.S: Data curation, review & editing;

T.S.H: Data curation, review & editing;

S.A: Data curation, review & editing;

R.R: Data curation, review & editing;

D.M: Data curation, review & editing;

M.S: Data curation, review & editing;

L.A: Data curation, review & editing;

A.U.B: Conceptualization, Supervision, Writing -review and editing;

C.F: Data curation, Writing - review & editing;

F.P: Conceptualization, Supervision, Writing -review and editing

## Data availability

The analysis pipeline is available at https://clinicalmpm.github.io/, including the sequence configuration for Siemens PRISMA scanners. MRI data from this study cannot be shared publicly due to constraints from the European General Data Protection Regulation and its implementation into German laws and required consent from participants.

