## Supplementary material for "A quantitative multi-parameter mapping protocol standardized for clinical research in autoimmune neuroinflammatory diseases with white matter abnormalities"

### Supplementary Figures

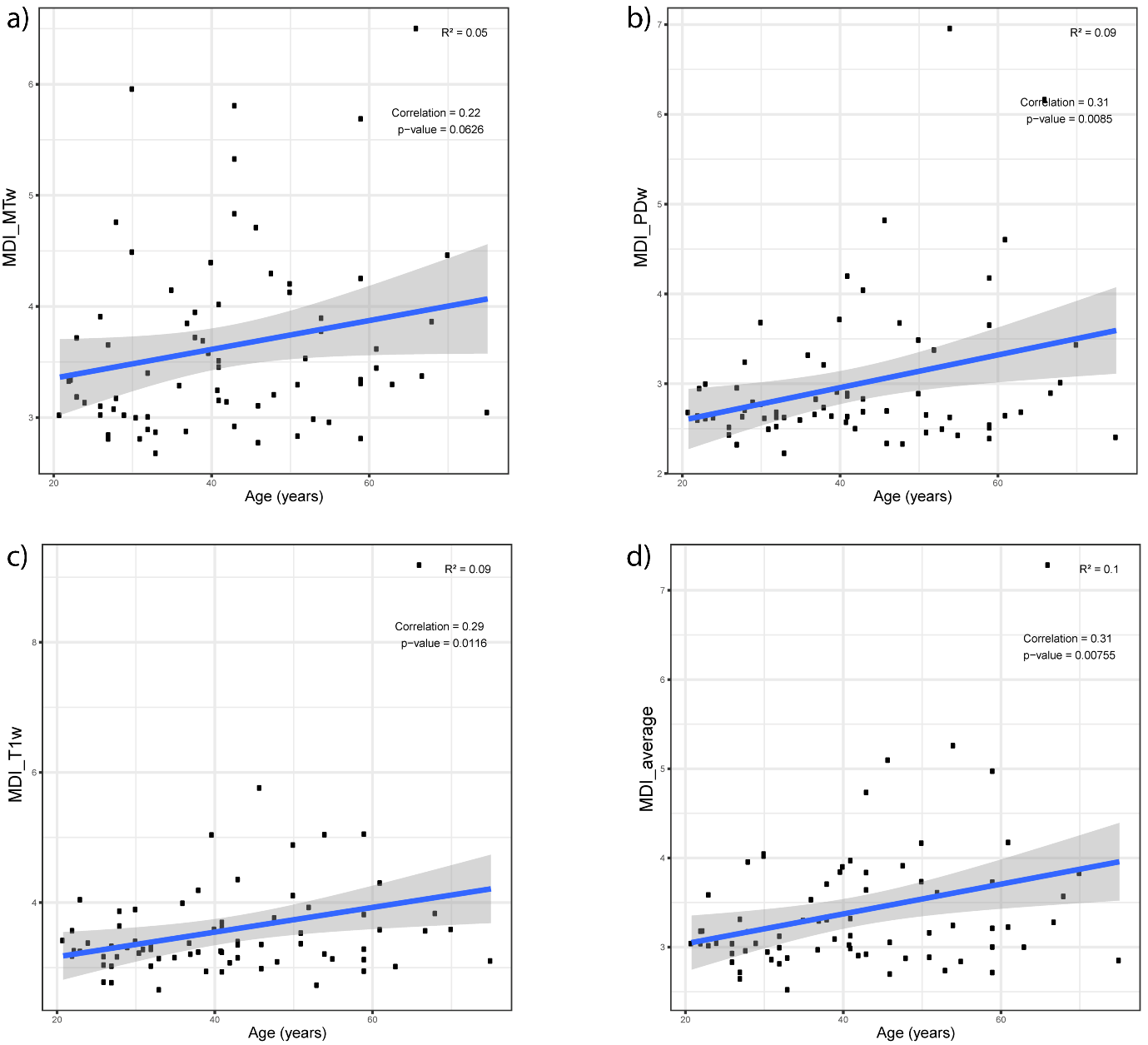

**Supplementary Figure S1.** **Motion degradation index correlation with age**. Scatterplots show correlation of Motion Degradation Index (MDI) with age for each of the raw average echo (MTw (a), PDw (b), T1w (c)) and the averaged sum of the respective MDI (d).

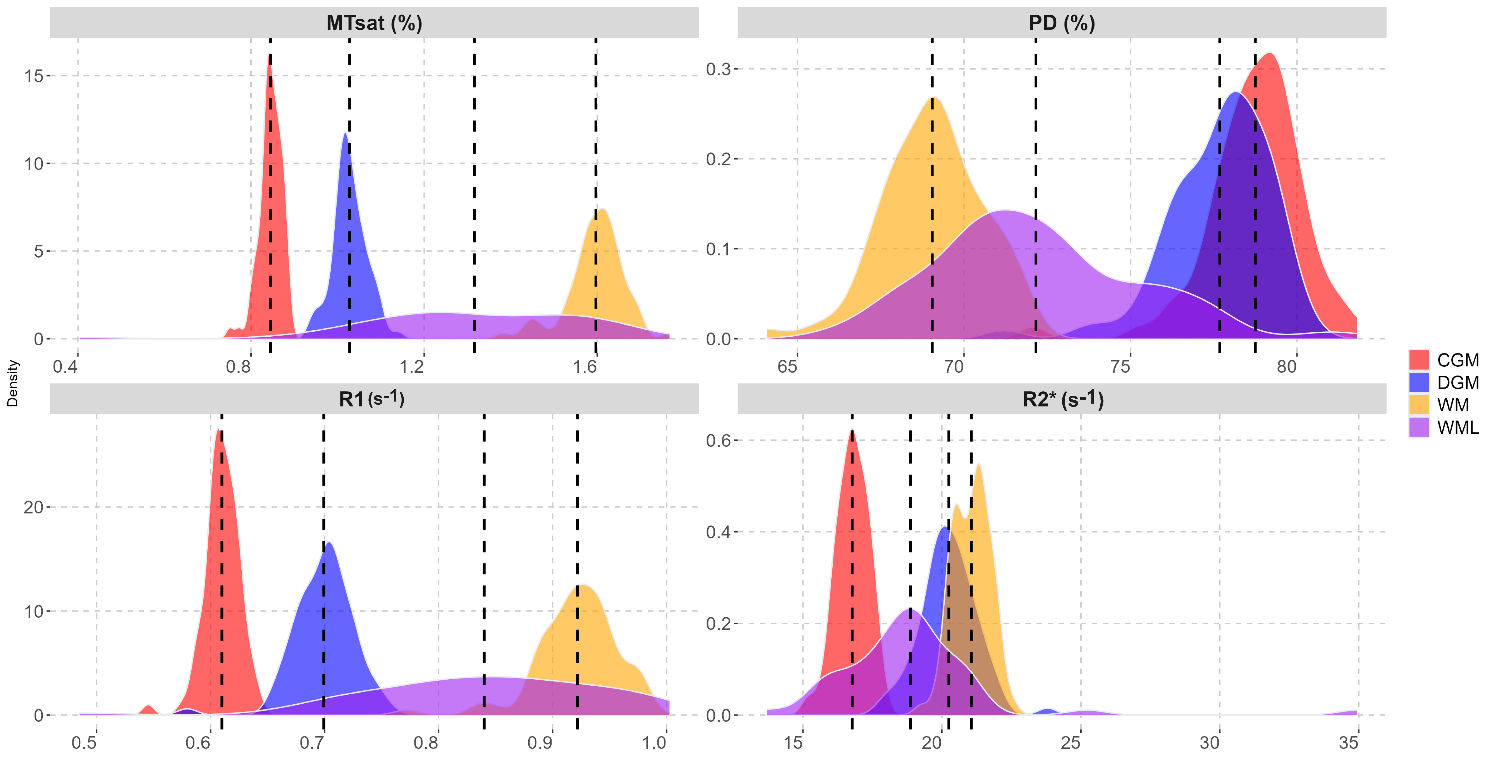

**Supplementary Figure S2.** **MPM values distribution**. Histograms of median MPM values distribution across participants on entire data acquired in white matter (WM, yellow), white matter lesions (WML, purple), cortical grey matter (CGM, red), deep grey matter (DGM, blue). Dashed vertical lines represent ROI median.

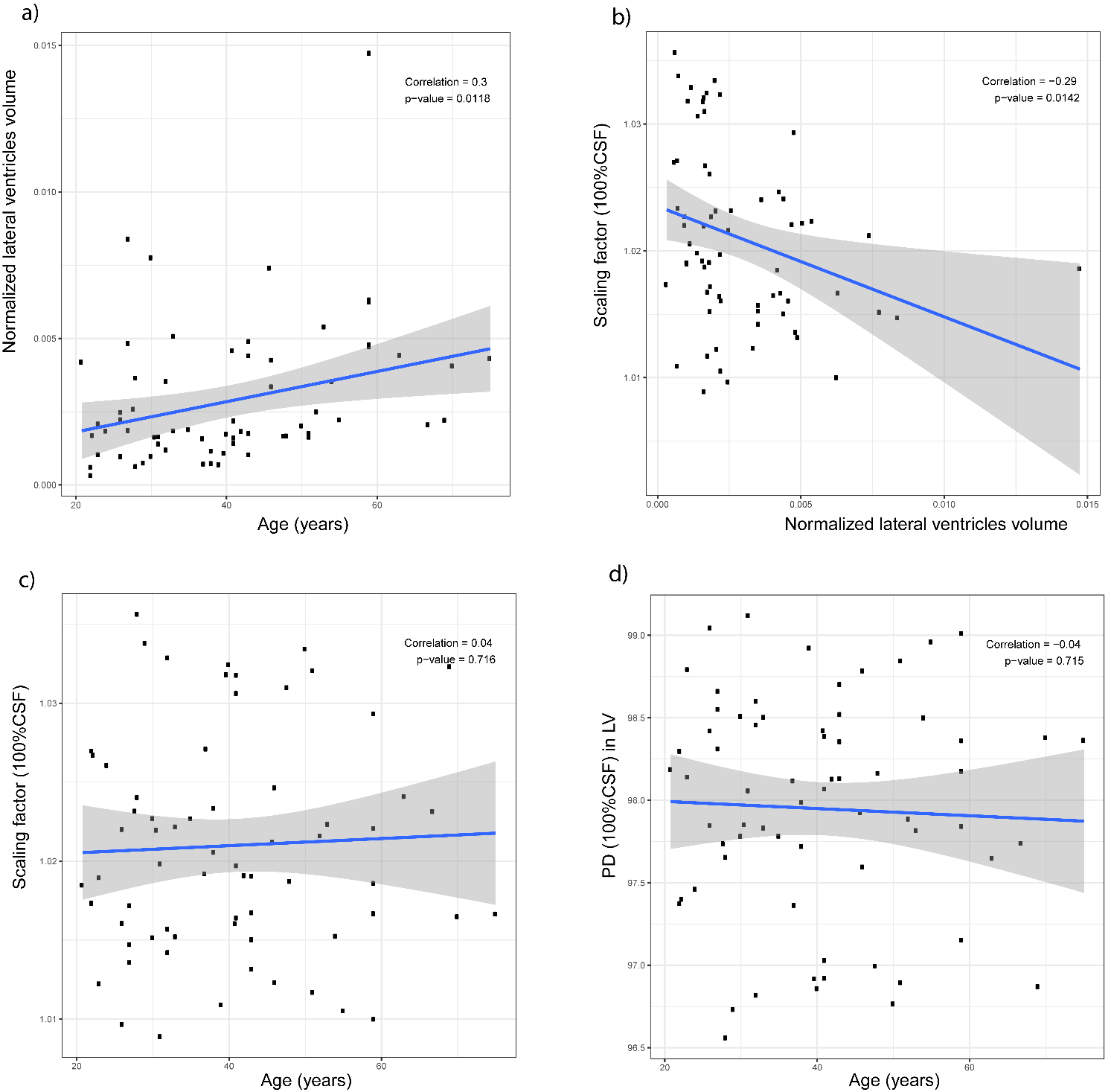

**Supplementary Figure S3.** **Calibration confounders.** Scatterplots showing correlation between normalized volume of lateral ventricles (LV) against age (a), between scaling factor for the calibration using CSF and normalized volume (b), between scaling factor and age (c), between PD and age (d).

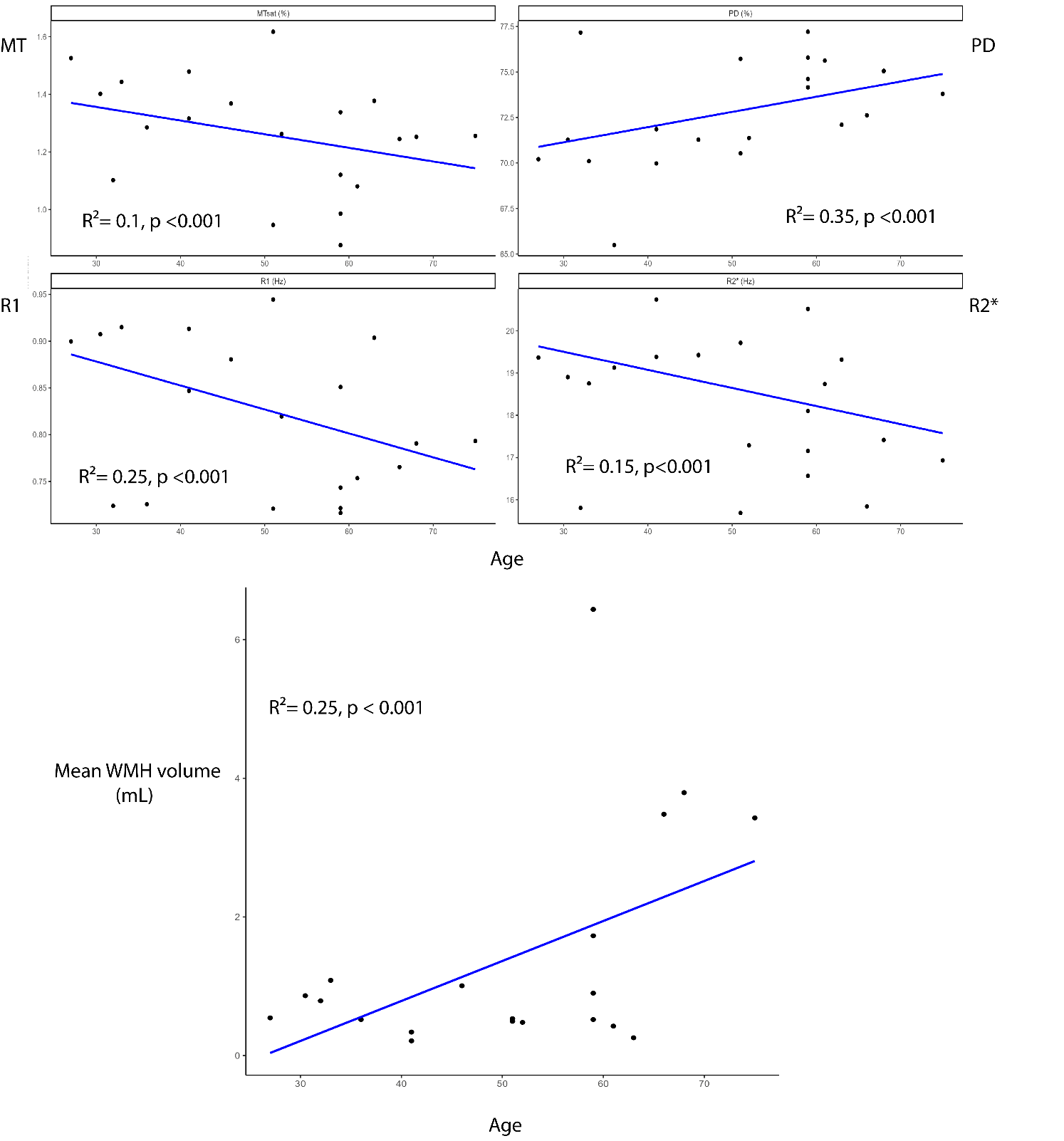

**Supplementary Figure S4. MPM scatterplots against age in lesions**. Scatterplots against age of median MPM values and volume in white matter hyperintensities (WMH, n=20 participants with lesion volume > 0.2mL).

A)
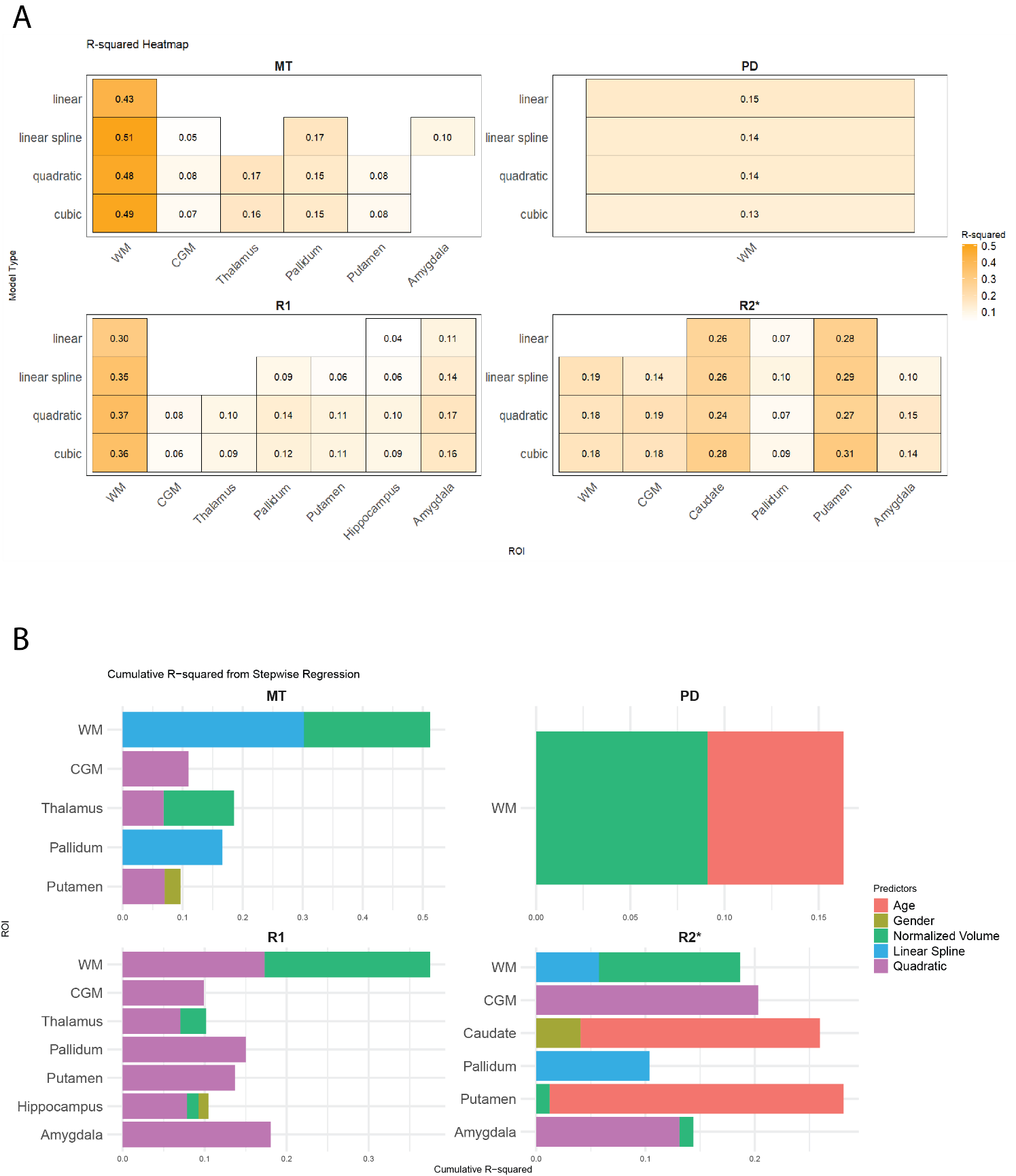

B)
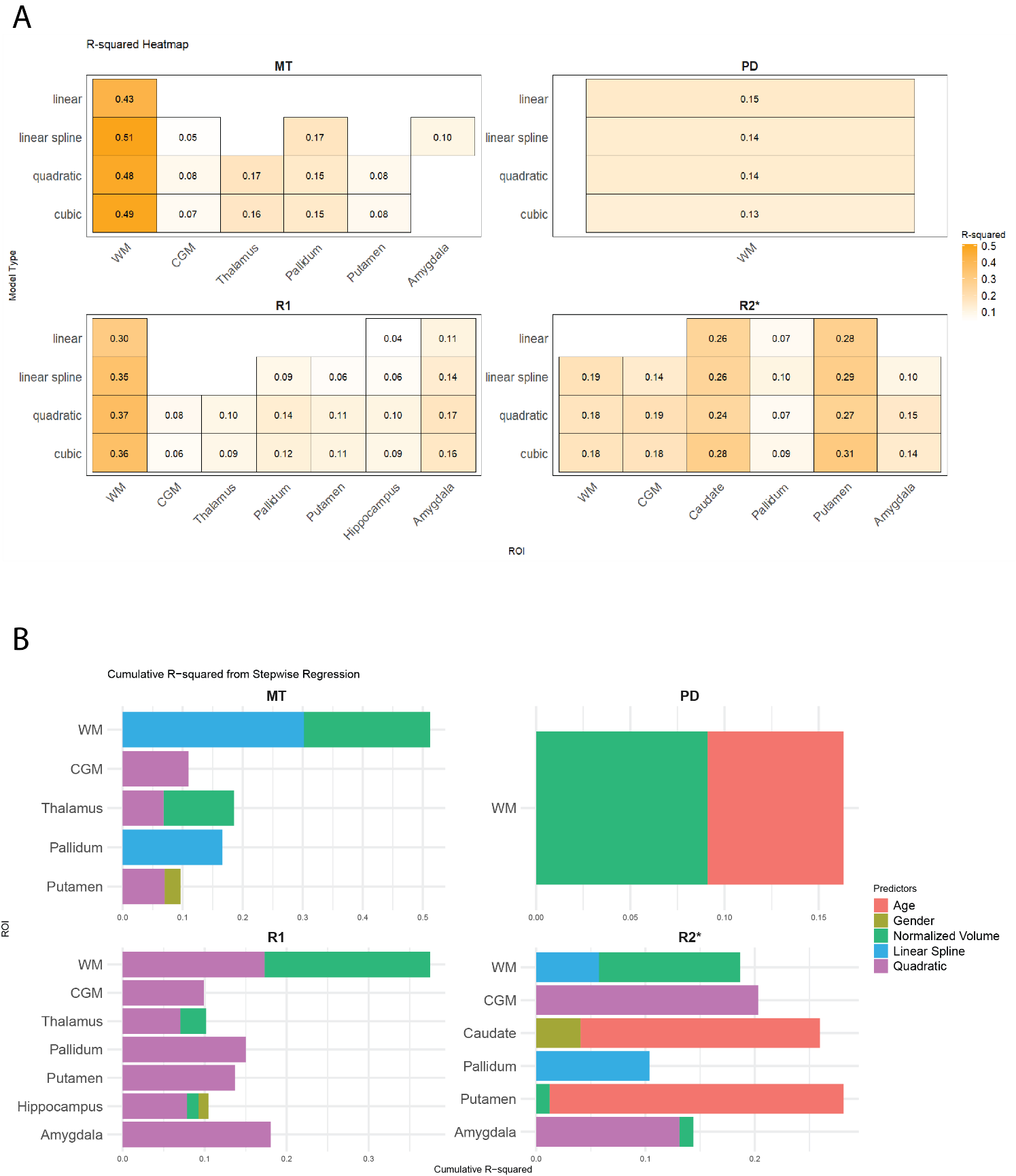

**Supplementary Figure S5:** Regression results of median MPM values against age.

**(A).** Adjusted R² comparison of regression models shown as heat maps for each quantitative parameter across ROIs. Only models presenting a significant linear, quadratic or cubic association to age are shown (lm() function in R). Exponential functions did not yield significant results and are therefore omitted.

**(B).** Summary of selected models with bar plots represent the cumulative R² contribution of each predictor. Only significant associations are shown.

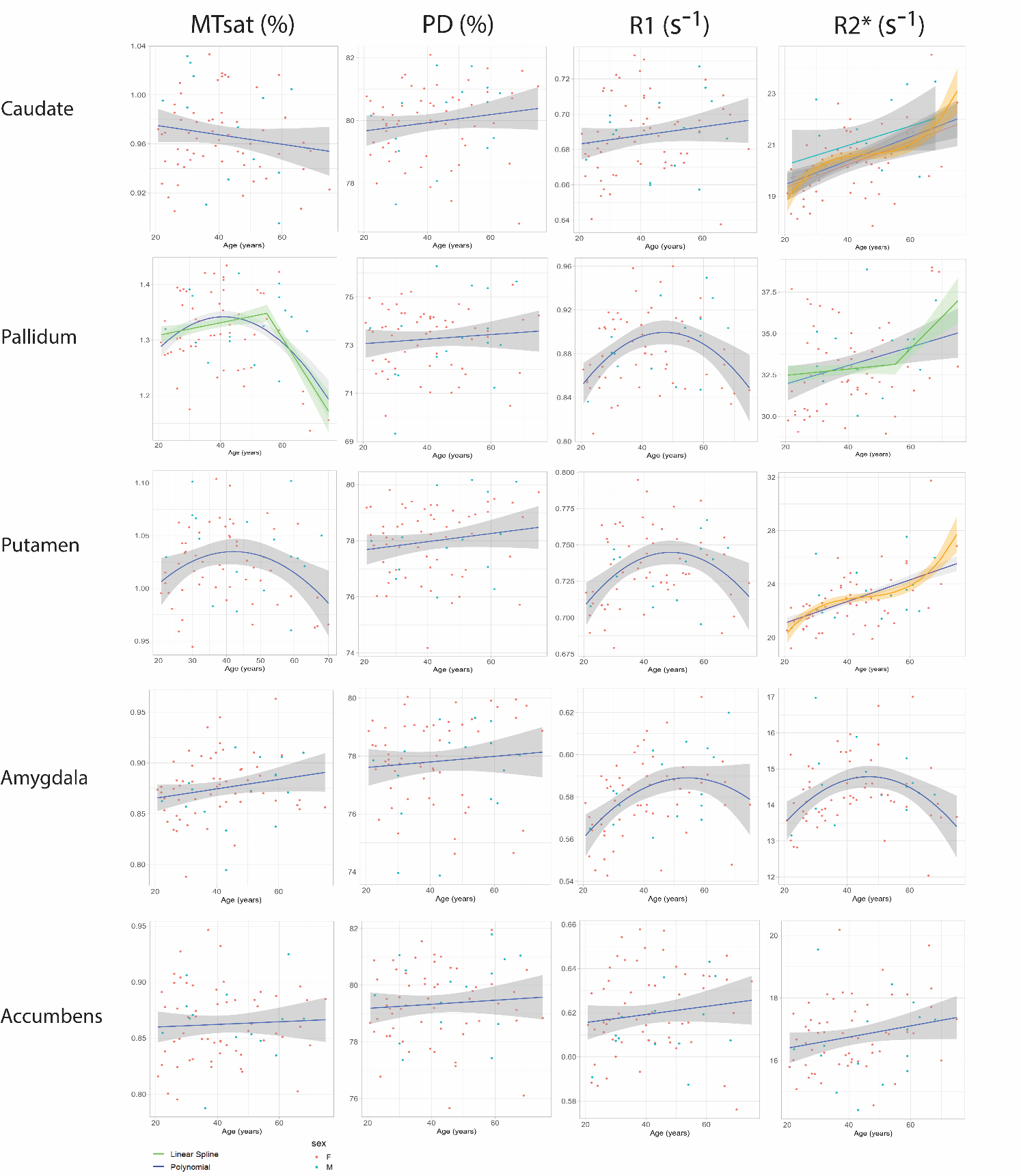

**Supplementary Figure S6.** Scatterplots and model fitting trajectories for non-described ROIs. Orange curve displays the cubic model which performed equally well but not significantly better than a linear regression in caudate nucleus and putamen.

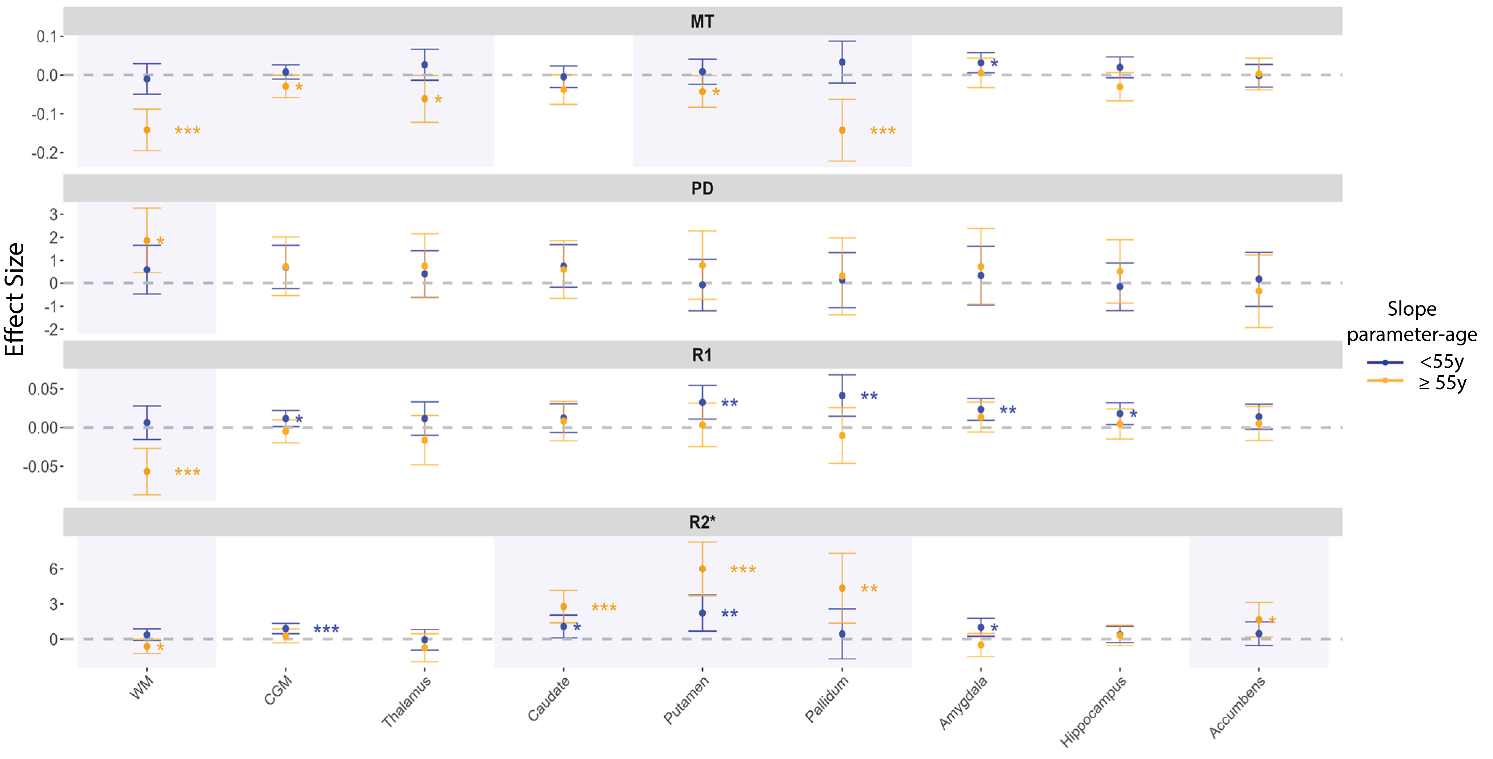

**Supplementary Figure S7.** Representation of linear spline slopes “parameter-age” before and after the cut-off of 55y across ROIs for MT, PD, R1, and R2^*^. Significance levels of parameter-age correlation are described with asterisks. Panel background is greyed out to help identify ROIs where the consideration of parameter variability might be important in individuals older than 55y (i.e. when linear age association is significant in the group of participants of age ≥55y).

### Supplementary Tables

| **Parameter** | | | | | |
| --- | --- | --- | --- | --- | --- |
| **MT** | | | | | |
| **ROI** | **Caudate** | **Pallidum** | **Putamen** | **Accumbens** | **Amygdala** |
| **Mean (SD)** | 0.97 (0.03) | 1.32 (0.07) | 1.03 (0.04) | 0.87 (0.03) | 0.88 (0.03) |
| **SD over voxels** | 0.16 | 0.15 | 0.15 | 0.12 | 0.14 |
| **Median (IQR)** | 0.97 (0.04) | 1.32 (0.10) | 1.02 (0.05) | 0.86 (0.04) | 0.88 (0.04) |
| **Intra-subject CoV** | 16.28 | 11.75 | 14.24 | 13.51 | 16.52 |
| **Inter-subject CoV** | 3.47 | 5.12 | 3.49 | 3.67 | 3.63 |
| **2nd-98th Percentile** | 0.91-1.04 | 1.17-1.41 | 0.97-1.11 | 0.81-0.94 | 0.8-0.94 |
| **Min-Max** | 0.9-1.04 | 1.14-1.44 | 0.96-1.11 | 0.8-0.95 | 0.8-0.95 |
| **PD** | | | | | |
| **ROI** | **Caudate** | **Pallidum** | **Putamen** | **Accumbens** | **Amygdala** |
| **Mean (SD)** | 79.7 (1.15) | 72.99 (1.36) | 77.69 (1.22) | 79.21 (1.34) | 77.76 (1.46) |
| **SD over voxels** | 2.71 | 3.03 | 2.42 | 2.54 | 2.92 |
| **Median (IQR)** | 79.95 (1.53) | 73.27 (1.99) | 77.99 (2.05) | 79.34 (1.93) | 77.82 (2.12) |
| **Intra-subject CoV** | 3.4 | 4.16 | 3.12 | 3.21 | 3.76 |
| **Inter-subject CoV** | 1.44 | 1.87 | 1.57 | 1.69 | 1.88 |
| **2nd-98th Percentile** | 77.07-81.57 | 70.11-75.46 | 75.54-79.74 | 76.38-81.43 | 74.23-79.83 |
| **Min-Max** | 76.69-81.67 | 69.06-75.64 | 74.32-79.88 | 75.89-81.7 | 74.04-79.96 |
| **R1** | | | | | |
| **ROI** | **Caudate** | **Pallidum** | **Putamen** | **Accumbens** | **Amygdala** |
| **Mean (SD)** | 0.69 (0.02) | 0.88 (0.03) | 0.73 (0.02) | 0.62 (0.02) | 0.59 (0.02) |
| **SD over voxels** | 0.07 | 0.06 | 0.06 | 0.05 | 0.06 |
| **Median (IQR)** | 0.69 (0.03) | 0.88 (0.05) | 0.73 (0.04) | 0.62 (0.03) | 0.58 (0.02) |
| **Intra-subject CoV** | 9.49 | 6.27 | 8.71 | 7.91 | 9.41 |
| **Inter-subject CoV** | 3.07 | 3.75 | 3.22 | 3.14 | 3.05 |
| **2nd-98th Percentile** | 0.65-0.73 | 0.82-0.95 | 0.69-0.78 | 0.59-0.66 | 0.55-0.62 |
| **Min-Max** | 0.65-0.73 | 0.8-0.97 | 0.68-0.79 | 0.57-0.66 | 0.55-0.63 |
| **R2*** | | | | | |
| **ROI** | **Caudate** | **Pallidum** | **Putamen** | **Accumbens** | **Amygdala** |
| **Mean (SD)** | 20.56 (1.31) | 32.84 (2.98) | 23.09 (2.21) | 16.94 (1.18) | 14.64 (0.93) |
| **SD over voxels** | 3.52 | 6.5 | 4.08 | 3.41 | 3.97 |
| **Median (IQR)** | 20.45 (1.39) | 33.17 (3.13) | 22.85 (2.39) | 16.79 (1.42) | 14.38 (1.12) |
| **Intra-subject CoV** | 17.10 | 19.45 | 17.40 | 20.13 | 27.13 |
| **Inter-subject CoV** | 6.36 | 9.06 | 9.58 | 6.95 | 6.36 |
| **2nd-98th Percentile** | 18.34-23.66 | 28.86-39.91 | 19.99-28.41 | 14.91-19.84 | 13.09-16.74 |
| **Min-Max** | 18.27-24.41 | 28.37-41.74 | 19.27-32.33 | 14.58-20.34 | 12.97-17.08 |
| **Volume** | | | | | |
| **ROI** | **Caudate** | **Pallidum** | **Putamen** | **Accumbens** | **Amygdala** |
| **Mean (SD) in mL** | 1.81 (0.3) | 1.41 (0.13) | 3.44 (0.36) | 0.4 (0.06) | 1.04 (0.21) |
| **ICV (mL)** | 2890 (256.05) | | | | |

**Supplementary Table S1.** Descriptive statistics table for non-described ROIs.

| **Reference study** | **ROI** | **Parameters** | | | |
| --- | --- | --- | --- | --- | --- |
| (Weiskopf et al., 2013)  N=5, age 24.2±1.6y  deviations to mean |  | **MT** | **PD** | **R1** | **R2*** |
|  | **WM** | 11.9% | 2.4% | 12% | 1.4% |
|  | **CGM** | 7% | 6.8% | 1.8% | 18.4% |
|  | **Caudate** | 16% | 3.6% | 1% | 13% |
|  | **Range across ROIs** | 7-16% | 2.4-6.8% | 1-12% | 1.4-13% |
| (Volz et al., 2012)  N=6, mean age 34y, range 25–71y |  |  |  |  |  |
|  | **WM** |  | 0.4% |  |  |
|  | **GM** |  | 2.8% |  |  |
|  | **Caudate** |  | 0.6% |  |  |
|  | **Putamen** |  | 2.6% |  |  |
|  | **Range across ROIs** |  | 0.4-2.8% |  |  |
| (Khalil et al., 2011)  N =35, age 36.7±13.7y  deviations to mean |  |  |  |  |  |
|  | **Pallidum** |  |  |  | 0.5% |
|  | **Putamen** |  |  |  | 2.6% |
|  | **Caudate** |  |  |  | 2.6% |
|  | **Thalamus** |  |  |  | 1.3% |
|  | **Range across ROIs** |  |  |  | 0.5-2.6% |
| (Callaghan et al., 2014)  N=138, age 46.6±21y  deviations to mean  (at 25y for MT and R1) |  | |  |  |  |
|  | **Thalamus** | 24.4% |  | 7.4% | 9.3% |
|  | **Hippocampus** | 10.8% |  | 3.8% |  |
|  | **Caudate** | 19.8% |  | 1.5% | 11.1% |
|  | **Amygdala** | 24% |  | 7.3% | 22% |
|  | **Putamen** | 6.2% |  |  | 11.3% |
|  | **Pallidum** |  |  |  | 16% |
|  | **Range across ROIs** | 6.2- 24.4% |  | 1.5-7.4% | 9.3-16% |
| (Leutritz et al., 2020)  N=5, age 32.4±6.0y  deviations to mean |  | |  |  |  |
|  | **WM** | 13.9% | 1.4% | 14.9% | 5.3% |
|  | **GM** | 8% | 2.2% | 1.6% | 11.2% |
|  | **Hippocampus** | 8% | 3.9% | 11.5% | 15.4% |
|  | **Caudate** | 9.1% | 3.7% | 6.2% | 9.4% |
|  | **Range across ROIs** | 8-14% | 1.4-4% | 1.6-15% | 5.3-15.4% |
| (Taubert et al., 2020)  N=966, (51% F),  age F: 60.7±9.4y, age M: 60±9.8y  deviations to mean |  |  | | | |
|  | **Putamen** | 6.4% | 0.9% |  | 7.6% |
|  | **Thalamus** | 7.0% | 3.4% |  | 8% |
|  | **Hippocampus** | 8.0% | 0.1% |  | 6.1% |
|  | **Range across ROIs** | 6.4-8% | 1-3% |  | 6-8% |
| (Hagiwara et al., 2018)  N=20, mean age 55.3y, age range 25–71  deviations to mean |  |  | |  |  |
|  | **WM** | 3.8% |  |  |  |
|  | **GM** | 0.1% |  |  |  |
|  | **Range across ROIs** | 0.1-3.8% |  |  |  |
| (Gracien et al., 2016)  N=11, age 43.6±11.2y  deviations to mean | **GM** |  | 2.4% | 12.7% |  |
|  | **WM** |  | 3% | 20.9% |  |
|  | **Caudate** |  | 0.6 | 8% |  |
|  | **Pallidum** |  | 1.9% | 10.2% |  |
|  | **Putamen** |  | 2.5% | 6.4% |  |
|  | **Thalamus** |  | 1.2% | 14% |  |
|  | **Range across ROIs** |  | 0.6-3% | 6.4-20.9% |  |
| (Lommers et al., 2019)  N = 36, age 45.9 ±12.5y  deviation to median |  | | | | |
|  | **WM** | 4.8% |  | 11.5% | 1.4% |
|  | **CGM** | 3.7% |  | 4.7% | 1.1% |
|  | **DGM** | 5.1% |  | 9.1% | 8.2% |
|  | **Range across ROIs** | 3.7-5.1% |  | 4.7-11.5% | 1.1-8.2% |

**Supplementary Table S2. Comparison of MPM values to literature.** Approximate MPM deviations to mean or median values found in literature across various ROIs when comparable. Range across ROIs represents minimum-maximum relative deviations observed.

| **ROI** | **T statistic** | **P value** |
| --- | --- | --- |
| WM | 1.71 | 0.091 |
| CGM | -3.47 | <0.001 |
| DGM | -1.98 | 0.051 |
| Amygdala | 1.82 | 0.074 |
| Hippocampus | 0.89 | 0.375 |
| Thalamus | -2.19 | 0.032 |
| Accumbens | -4.28 | <0.001 |
| Caudate | -0.39 | 0.699 |
| Pallidum | -1.61 | 0.11 |
| Putamen | -3.64 | <0.001 |
| Lateral ventricles | 2.59 | 0.012 |

**Supplementary Table S3:** Pearson correlation of normalized structure volume vs age. Structural volumes were normalized by taking respective volume divided by ICV.

| **Parameter** | **ROI** | **Predictors** | **Coefficient β** | **P-value** | **Confidence Interval (95%)** |
| --- | --- | --- | --- | --- | --- |
| **MT** | **Putamen** | **Selected model: quadratic fit (F=3.56, p=0.018)** | | | |
|  |  | **Intercept** | 1.020 | <0.001 | (1.011, 1.03) |
|  |  | **Age** | -0.040 | 0.272 | (-0.113, 0.033) |
|  |  | **Age²** | -0.096 | 0.008 | (-0.168, -0.026) |
|  |  | **Sex: F vs M** | 0.018 | 0.085 | (-0.003, 0.038) |
|  | **Pallidum** | **Selected model: linear spline (F=8.17, p<0.001)** | | | |
|  |  | **Intercept** | 1.309 | <0.001 | (1.28, 1.34) |
|  |  | **Age (slope before 55)** | 0.039 | 0.146 | (-0.014, 0.091) |
|  |  | **Age (slope after 55)** | -0.138 | 0.001 | (-0.214, -0.063) |
| **R1** | **Putamen** | **Selected model: quadratic fit (F=6.71, p=0.002)** | | | |
|  |  | **Intercept** | 0.734 | <0.001 | (0.729, 0.74) |
|  |  | **Age** | 0.040 | 0.088 | (-0.007, 0.088) |
|  |  | **Age²** | -0.076 | 0.002 | (-0.123, -0.029) |
|  | **Pallidum** | **Selected model: quadratic fit (F=7.36, p=0.001)** | | | |
|  |  | **Intercept** | 0.884 | <0.001 | (0.877, 0.892) |
|  |  | **Age** | 0.036 | 0.242 | (-0.026, 0.098) |
|  |  | **Age²** | -0.112 | 0.001 | (-0.174, -0.051) |
|  | **Amygdala** | **Selected model: quadratic fit (F=8.93, p<0.001)** | | | |
|  |  | **Intercept** | 0.580 | <0.001 | (0.577, 0.585) |
|  |  | **Age** | 0.056 | 0.001 | (0.024, 0.088) |
|  |  | **Age²** | -0.040 | 0.016 | (-0.073, -0.008) |
| **R2*** | **Caudate** | **Selected model: linear fit (F=13.62, p<0.001)** | | | |
|  |  | **Intercept** | 18.510 | <0.001 | (17.655, 19.365) |
|  |  | **Age** | 0.043 | <0.001 | (0.023, 0.063) |
|  |  | **Sex: F vs M** | 0.715 | 0.030 | (0.070, 1.360) |
|  | **Putamen** | **Selected model: linear fit (F=15.08, p<0.001)** | | | |
|  |  | **Intercept** | 15.523 | <0.001 | (10.091, 20.957) |
|  |  | **Age** | 0.093 | <0.001 | (0.059, 0.128) |
|  |  | **Normalized Volume** | 991.789 | 0.141 | (-336.901, 2320.480) |
|  | **Pallidum** | **Selected model: linear spline (F=5.16, p=0.008)** | | | |
|  |  | **Intercept** | 32.492 | <0.001 | (31.315, 33.669) |
|  |  | **Age (slope before 55)** | 0.653 | 0.526 | (-1.394, 2.7) |
|  |  | **Age (slope after 55)** | 4.481 | 0.002 | (1.683, 7.279) |
|  | **Amygdala** | **Selected model: quadratic fit (F=5.03, p=0.003)** | | | |
|  |  | **Intercept** | 13.046 | <0.001 | (11.165, 14.928) |
|  |  | **Age** | 0.657 | 0.452 | (-1.078, 2.392) |
|  |  | **Age²** | -3.059 | 0.001 | (-4.785, -1.335) |
|  |  | **Normalized Volume** | 1168.943 | 0.158 | (-465.516,2803.402) |

**Supplementary Table S4.** Model selection results for non-described ROIs. Regression coefficients β and 95% confidence interval are reported for each independent variable.

| **Parameter** | **ROI** | **Model predictors** | **Coefficient ß** | **Confidence Interval (95%)** | **P-value** |
| --- | --- | --- | --- | --- | --- |
| PD | Accumbens | Selected model: Linear (F=1.53, p=0.224) | |  |  |
|  |  | Intercept | 81.594 | (78.308,84.881) | <0.001 |
|  |  | Normalized Volume | -5163.607 | (-11522.80 ,1195.585) | 0.109 |
|  |  | Age | -0.003 | (-0.028,0.022) | 0.795 |
|  | Amygdala | Linear (F=1.97, p=0.148) | |  |  |
|  |  | Intercept | 74.573 | (71.271, 77.875) | <0.001 |
|  |  | Normalized Volume | 2614.197 | (-260.347,5488.741) | 0.074 |
|  |  | Age | 0.006 | (-0.018,0.031) | 0.610 |
|  | Caudate | Linear (F=5.84, p=0.004) | |  |  |
|  |  | Intercept | 76.997 | (75.253,78.74) | <0.001 |
|  |  | Age | 0.015 | (-0.003,0.033) | 0.095 |
|  |  | Normalized Volume | 1251.986 | (445.931,2058.041) | 0.003 |
|  | CGM | Linear (F=4.66, p=0.034) | |  |  |
|  |  | Intercept | 78.006 | (77.223,78.789) | <0.001 |
|  |  | Age | 0.019 | (0.001,0.036) | 0.034 |
|  | DGM | Linear (F=5.04, p=0.009) | |  |  |
|  |  | Intercept | 71.758 | (67.547,75.969) | <0.001 |
|  |  | Age | 0.027 | (0.007,0.048) | 0.009 |
|  |  | Normalized Volume | 303.991 | (64.016,543.966) | 0.014 |
|  | Hippocampus | Linear (F=5.76, p=0.005) | |  |  |
|  |  | Intercept | 75.499 | (73.321,77.677) | <0.001 |
|  |  | Normalized Volume | 1273.000 | (523.589,2062.719) | 0.002 |
|  |  | Age | 0.006 | (-0.014,0.027) | 0.532 |
|  | Pallidum | Linear (F=0.65, p=0.425) | |  |  |
|  |  | Intercept | 72.87 | (71.835,73.906) | <0.001 |
|  |  | Age | 0.01 | (-0.014,0.033) | 0.425 |
|  | Putamen | Linear (F=1.89, p=0.173) | |  |  |
|  |  | Intercept | 77.382 | (76.449,78.315) | <0.001 |
|  |  | Age | 0.015 | (-0.007,0.036) | 0.173 |
|  | Thalamus | Linear (F=2.89, p=0.093) | |  |  |
|  |  | Intercept | 75.9 | (75.027,76.773) | <0.001 |
|  |  | Age | 0.017 | (-0.003,0.036) | 0.093 |
|  | WM | Linear (F=8.02, p=0.001) | |  |  |
|  |  | Intercept | 76.119 | (70.538,81.701) | <0.001 |
|  |  | Age | 0.033 | (0.013,0.054) | 0.002 |
|  |  | Normalized Volume | -46.328 | (-77.615, -15.04) | 0.004 |
|  | Lateral ventricles | Age | 7.44e-05 | (-4.92e-05, 1.98e-04) | 0.234 |
|  |  | Normalized Volume | -1.002 | (-1.72, -0.28) | 0.007 |

**Supplementary Table S5.** Fitting results of PD against age. All significant and non-significant age associations across ROIs are reported.

### Supporting information figure captions

**Sup. Fig. S1. Motion degradation index correlation with age.** Scatterplots show correlation of Motion Degradation Index (MDI) with age for each of the raw average echo (MTw (a), PDw (b), T1w (c)) and the averaged sum of the respective MDI (d).

**Sup. Fig. S2. MPM values distribution.** Histograms of median MPM values distribution across participants on entire data acquired in white matter (WM, yellow), white matter lesions (WML, purple), cortical grey matter (CGM, red), deep grey matter (DGM, blue). Dashed vertical lines represent ROI median.

**Sup. Fig. S3.** **Calibration confounders.** Scatterplots showing correlation between normalized volume of lateral ventricles (LV) against age (a), between scaling factor for the calibration using CSF and normalized volume (b), between scaling factor and age (c), between PD and age (d).

**Sup. Fig. S4.** **MPM scatterplots against age in lesions**. Scatterplots against age of median MPM values and volume in white matter hyperintensities (WMH, n=20 participants with lesion volume > 0.2mL).

**Sup. Fig. S5: Regression results of median MPM values against age.**

**(A).** Adjusted R² comparison of regression models shown as heat maps for each quantitative parameter across ROIs. Only models presenting a significant linear, quadratic or cubic association to age are shown (lm() function in R). Exponential functions did not yield significant results and are therefore omitted.

**(B).** Summary of selected models with bar plots represent the cumulative R² contribution of each predictor. Only significant associations are shown.

**Sup. Fig. S6.** **Scatterplots and model fitting trajectories for non-described ROIs.** Orange curve displays the cubic model which performed equally well but not significantly better than a linear regression in caudate nucleus and putamen.

**Sup. Fig. S7.** **Representation of linear spline slopes “parameter-age” before and after the cut-off of 55y across ROIs for MT, PD, R1, and R2^*^.** Significance levels of parameter-age correlation are described with asterisks. Panel background is greyed out to help identify ROIs where the consideration of parameter variability might be important in individuals older than 55y (i.e. when linear age association is significant in the group of participants of age ≥55y).

**Sup. Table S1. Descriptive statistics table for non-described ROIs**.

**Sup. Table S2. Comparison of MPM values to literature.** Approximate MPM deviations to mean or median values found in literature across various ROIs when comparable. Range across ROIs represents minimum-maximum relative deviations observed.

**Sup. Table S3: Pearson correlation of normalized structure volume vs age**. Structural volumes were normalized by taking respective volume divided by ICV.

**Sup. Table S4. Model selection results for non-described ROIs.** Regression coefficients β and 95% confidence intervals are reported for each independent variable.

**Sup. Table S5. Fitting results of PD against age**. All significant and non-significant age associations across ROIs are reported.
